# Stochastic Chaos in Influenza Data – An Application of Topological Methods

**DOI:** 10.1101/2025.07.09.25331183

**Authors:** Carlos Pedro Gonçalves, Carlos Rouco

## Abstract

Empirical methods of chaos theory have been applied to epidemiological data, uncovering evidence of chaos. In the current work, we apply, to the weekly share of positive tests for influenza in the Northern Hemisphere, an empirical methodology for studying stochastic chaos using topological data analysis methods combining chaos theory, topological machine learning, and nonlinear time series analysis for attractor reconstruction and decomposition to decompose a stochastic chaotic dynamics down to the independent and identically distributed (IID) noise. A stochastic chaotic attractor is found for the epidemiological series comprised of an interacting chaotic dynamics and a nonlinear stochastic component with nonstationary variance; the full dynamics is researched down to the IID noise, and the resulting estimated model is used to analyze major risk metrics, including probability dynamics and changing volatility. The implications for epidemiological empirical studies applying chaos theory to epidemiological data are discussed.

## 1. Introduction

Chaos theory has been applied to epidemiology in both theoretical models and in empirical studies, with markers of chaos having been found in different epidemiological series [1–10]. This last evidence leads to the possibility of using chaos theory’s empirical methods for both researching basic epidemiological processes and the dynamics of epidemiological statistics with impact in epidemiological risk analysis and healthcare management.

The dynamics of target epidemiological variables linked to viruses result from different complex processes including, among others, environmental factors, epidemiological vectors, mutation rates, immune response, people’s interactions, vaccination and healthcare policies as well as testing processes that lead to the identification of positive cases for a viral infection, in this way, the dynamics of key epidemiological statistical variables like positive cases for viral infections are the result of complex dynamics, including feedback loops between such statistics and healthcare responses and policies which further increase the complexity of this dynamics, since healthcare authorities can use these statistics as a way to plan and implement adaptive responses to viral spread which has consequences in the epidemiological dynamics itself [5,6,10].

In this context, emergent chaotic attractors found in time series of target epidemiological variables constitute an example of complex nonlinear dynamics with relevance for healthcare planning and risk management, namely, chaotic attractors, while unpredictable in the long run due to an exponential amplification of small deviations also, have topological signatures that allow for prediction within bounds [5,6,11].

While the identification of chaotic attractors in data and the study of these attractors in terms of their deterministic components through noise filtering is a critical point in empirical research on chaos [12]. In complex systems, stochastic chaos can occur with complex patterns in which the nonlinear deterministic component can interact with nonlinear stochastic processes with consequences for the analysis of the systems’ dynamics [13–16]. In multicausal contexts involving complex systems’ dynamics in coevolutionary scenarios, chaos is usually stochastic rather than deterministic.

The difference between deterministic chaos and stochastic chaos is that, while deterministic chaos deals with a nonlinear deterministic dynamics with random-like trajectories sensitive to small deviations, stochastic chaos deals with an open system’s dynamics that involves a deterministic component that is chaotic and a stochastic component that affects the system’s dynamics and plays a role in it [5,6,13–16].

In this sense, in stochastic chaos, markers of chaotic attractors can be found but these attractors are affected by dynamical noise which can, itself, have complex nonlinear features, indeed, stochastic chaos is a stochastic process with a nonlinear deterministic component that is chaotic, thus, the process typically leads to noisy attractors that can exhibit major markers of chaos, including the sensitive dependence to small fluctuations and the fractal signatures [14–16].

In complex systems in general, and in the epidemiological context in particular, chaos, as stated, is typically stochastic [5, 6]. In this sense, the employment of empirical methodologies for studying chaotic attractors needs to study both reconstructed attractors and the stochastic component in tandem, decomposing, if possible, the full process down to independent and identically distributed (IID) noise, since complex stochastic processes can occur in tandem with the chaotic component and even be influenced by it in terms of feedback loops, as we will see in the present work for the case of the influenza data.

An empirical topological methodology, employing topological data analysis methods involving machine learning and attractor reconstruction, coupled with nonlinear time series analysis methods, can actually be employed to produce a decomposition between the deterministic chaotic component and the stochastic component, allowing for the decomposition of the system’s dynamics down to IID noise [13], which, in turn, allows for probabilistic analysis of the main dynamics.

In the present work, we illustrate the application of such a methodology to an influenza time, series from the FluNet dataset, by the World Health Organization, processed by *Our World In Data* in the indicator *share of positive tests – all types of surveillance*, which involves calculating a share of positive tests to any influenza strain for different regions and countries, the region we will be researching is the Northern Hemisphere, which provides for an aggregate indicator giving an overall picture of influenza dynamics in that hemisphere.

The use of this aggregate indicator reduces possible effects of testing frequencies and provides for an example of an epidemiological surveillance variable and how complex dynamics can occur for such variables.

We find that the above influenza series is, actually, characterized by a two-dimensional chaotic attractor affected by nonlinear dynamical noise, namely, we find the presence of chaos that is near a bifurcation point between a chaotic regime and periodic or quasiperiodic dynamics (*onset of chaos*), characterized by long-range persistence, along with a type of nonlinear stochastic process with nonstationary variance called generalized autoregressive conditional heteroskedasticity (GARCH), with standardized residuals that are found to be IID and characterized by a Student’s *t* distribution.

The underlying chaotic component accounts for around 78% of the variability of the share of positive tests for influenza in the Northern Hemisphere, with an *R*^2^ of 78.33% and an explained variance of 78.34%. There is also a feedback from the chaotic component to the nonlinear stochastic process.

Conjointly, the full deterministic dynamics which includes the chaos plus GARCH components account for around 97% of the above target epidemiological series, with an *R*^2^ of 97.14% and an explained variance of 97.49%.

We use the estimated model to calculate and characterize the probability dynamics and changing volatility for the target epidemiological series. We also show how the COVID-19 period affected the dynamics in the form of a contraction in the dynamical mean and variance of the full stochastic chaotic process.

While applied to this specific epidemiological series, the methodology is generalizable for any epidemiological time series as a way to uncover possible attractors and use the attractor decomposition method to research the dynamics down to the IID noise component with subsequent calculation of relevant epidemiological metrics.

In section 2, we review the main concepts and methods, addressing the main methodological steps employed. In section 3, we apply the methodology to our target series and, in section 4, we discuss the main findings.

## 2. Materials and Methods

Chaos theory developed within the research field of nonlinear dynamics researching the processes through which both low-dimensional and high-dimensional nonlinear deterministic dynamical systems could generate random-like dynamics [11,16,17]. The theory then expanded to address the interaction of such systems with noise, leading to the research line of stochastic chaos [13–16].

Deterministic chaos occurs in nonlinear dynamics as a bounded nonperiodic dynamics for which any small deviation in the state of the system is amplified with time at an exponential rate [11]. Two types of chaos have been researched within nonlinear dynamics, one is dissipative chaos the other is conservative chaos. Considering the geometric space spanned by a dynamical system’s degrees of freedom, with each degree of freedom corresponding to a dynamical variable describing the system’s dynamics, conservative chaos has a dynamics that conserves volumes in phase space, while dissipative chaos has an expansion and contraction dynamics in a bounded region leading to fractal attractors technically known as strange attractors [11,17].

In the context of complex systems affected by noise, usually chaos is of the latter kind, that is, dissipative with the noise itself leading to a dissipative dynamics, while conservative chaos corresponds to systems that can be described by a Hamiltonian formalism [11,16,17].

As we will find, in the present work, the empirical evidence for influenza is favorable to a type of emergent dynamics that is low-dimensional and weakly dissipative, indicating the possibility of a form of low-dimensional Hamiltonian chaos with a weakly dissipative component and a nonlinear noise process, leading to a weakly chaotic strange attractor very near a transition point between a nonchaotic dynamics and chaotic dynamics, that is, near the onset of chaos. This point will be discussed throughout the work and in the conclusions in connection to Hamiltonian models in epidemiology.

The randomness associated with chaotic attractors is linked to two major common factors. The first factor is the bounded dynamics; that is, the dynamics within a chaotic attractor is confined to a bounded region in the geometric space spanned by the dynamical system’s number of dynamical variables (the number of degrees of freedom). The second factor is the dynamical instability associated with the sensitive dependence upon initial conditions, which is linked to the exponential rate of divergence of small neighborhoods of any point in the attractor [11,16,17].

A chaotic attractor is thus stable, in the sense that it is invariant under the system’s dynamics and attracts nearby orbits, however, it also has an internal dynamical instability linked to the rate at which small deviations are exponentially amplified. This last point also means that small approximations to measured dynamical variables, and finite point rounding of floating-point numbers in computers render the dynamics unpredictable in the long run when dealing with chaotic orbits [11,16,17].

While this bounded, random-like nonperiodic dynamics with sensitive dependence upon initial conditions might seem to be unpredictable there is a topological structure in chaos associated with a skeleton of unstable periodic orbits that leads to regularities in the form of localized recurrences that allow for prediction within the limits set by the exponential rate of divergence, which can be measured mathematically by the largest Lyapunov exponent [11,16,17].

From a topological standpoint, a chaotic dynamics can stay for some time close to a periodic orbit until the exponential divergence sets in, leading it close to another periodic orbit. In this way, the structure of recurrences linked to the skeleton of unstable periodic orbits is a fundamental part of both the theoretical and empirical methods of chaos theory, which effectively use topological analysis as a way for addressing chaos in empirical data [5,6,11,17,18].

There are two major markers of chaos in empirical data; one is a positive largest Lyapunov exponent, which corresponds to the rate of exponential divergence of small neighborhoods under the phase space dynamics or, more simply put, the strength of the butterfly effect [11].

The second marker is the presence of exploitable topological information for the prediction of the system, taking advantage of the recurrence structure in chaos [5,6,11,17,18]. This last point plays a critical role in the reconstruction of a multidimensional chaotic attractor from a one-dimensional signal and has been used in both epidemiological series and other empirical applications of chaos theory [5–8,13,18,19].

The basic theory for how, given a time series, one can reconstruct an underlying attractor is covered by Takens’ work and has become a fundamental part of the empirical methods of chaos theory [11,19].

The main point is that given a time series signal generated by a multidimensional chaotic attractor, one can reconstruct that attractor by embedding the time series in multidimensional Euclidean space using an appropriate time delay and dimension, such that, even without knowing the basic equations, one can study the reconstructed trajectory in the above multidimensional space, a study that involves the application of topological analysis methods that formed an early part of chaos theory’s empirical methods and that have become integrated more recently into the wider field of topological data analysis, with the delay embedding becoming a critical step in the methodological approach when dealing with a system’s dynamics [5, 6, 8, 13].

Given a multidimensional dynamical state vector *u*, and a time series of observations that is a smooth function of *u*:

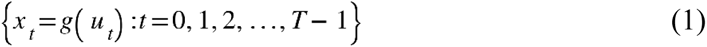

There is a dimension *d* and a delay lag *h* such that the main properties of the trajectory of *u* in phase space can be recovered from the time series by constructing the following vector in *d*-dimensional Euclidean space [11]:

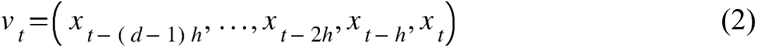

In this way, even without knowing the true state vector *u* and the main dynamical equations, with just a one-dimensional signal one can reconstruct the underlying system’s dynamics with its major geometric and topological properties through the above method as addressed by Takens [11,19].

The main issue is then that of finding the two embedding parameters, the dimension *d* and lag *h*, for which the underlying attractor’s topological structure is uncovered.

Topological data analysis, using topological machine learning, offers a way to directly link these embedding parameters and the reconstructed trajectory to the predictability of the target series from the topological signatures in the reconstructed phase space dynamics, such methods that have been employed in different contexts, ranging from epidemiology to sunspot dynamics, have been shown to be both effective on attractors and on bifurcations when attractor stability changes leading to a transient period followed by a new attractor as was shown in [5] for COVID-19, this also makes them effective in contexts with long-range dependence including fractal and multifractal scaling in the original signal and chaos affected by turbulent noise with nonstationary variance [6,8,13].

The main approach is to use topological machine learning as a search algorithm for optimal embedding parameters within a range of alternative values, that is, to use topological machine learning to find the embedding for which an adaptive artificially intelligent agent with a topological machine learning unit, operating on local patterns in the reconstructed phase space trajectory, is capable of extracting the most topological information in the prediction of the target series. Formally, the method involves using machine learning for extraction of local topological patterns in order to predict the target series, using different embedding parameters [5,6,8,13].

In order to account for possible bifurcations or the presence of dynamical noise leading to dynamical regimes associated with stochastic chaos, one should use a sliding window for relearning, allowing the artificial agent equipped with a topological machine learning unit to adapt to the local dynamical patterns of an attractor.

The main reason for using a sliding window, in the case of possible chaotic dynamics, is linked to the unstable periodic orbits’ skeleton that forms the topological order underlying a chaotic attractor [11,17,18], in this way, a window that is too long will capture different unstable cycles to which the attractor comes close and that are followed for a while until the exponential divergence associated with chaos leads it away from that neighborhood. The capturing of different cycles may lead to a reduction in the ability to capture a local topological structure.

Thus, using the full dataset for learning could fail in capturing the local topological dynamics associated with the recurrence structure of a chaotic attractor, which makes the sliding window a necessity when dealing with the search for embeddings that capture the underlying attractor’s topological structure.

In the current work, we will use a *k*-nearest neighbors’ learning unit. This type of unit is useful when dealing with turbulent data and can be used to make a bridge to the subsequent topological analysis of the phase space reconstructed trajectory using *k*-nearest neighbors’ graphs.

Therefore, given an embedding of the trajectory from a time series of observations, for a given dimension *d* and lag *h*, the training data is given by the sliding windows comprised of the ordered pairs of phase space vectors *v* and time series signal *x*:

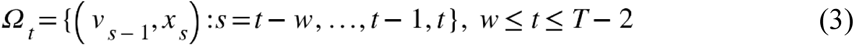

Given the training window data, an adaptive topological agent with a *k*-nearest neighbors’ learning unit is, thus, trained to predict the target series *x* given the previous phase point *v*, then, the adaptive topological agent is used to predict the first value of the series outside the training window, given the previous phase point, formally, we have the conditional expectation produced by the agent, trained on the training set given in equation (3):

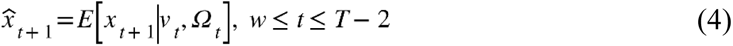

The process is continued until the end of the series so that the last value to be predicted is the series value at *T* – 1. The agent’s prediction performance is then calculated and recorded. This process is repeated for different embedding parameter values and the embedding leading to the best prediction performance is selected with the corresponding reconstructed attractor being researched on its main dynamics including the identification for markers of chaos.

The process, thus, involves a grid search from a set of alternative embedding dimensions and lags for the alternative where the topological patterns are best captured by the adaptive topological agent; in this sense, one is trying to identify, from the alternative embeddings, the one where there is the highest exploitable topological information.

After selecting an embedding, we provide the main performance metrics for the adaptive topological agent, including the *R*^2^ score, which we will use as a performance metric to select the embedding; the explained variance; the correlation between the agent’s predictions and the observed series; the root mean squared error (RMSE); and the relative error measured by the quotient of the RMSE by the total data amplitude.

We also study the pattern of the *R*^2^ score for the different lags and optimal dimensions obtained for each lag as a first topological data analysis method in order to find how the optimal dimension changes with the lag and the pattern of performance with different embedding dimensions.

Besides the above metrics, we also employ, on the final reconstructed phase space trajectory, Eckmann *et al.*’s method for the Lyapunov spectrum estimation [20]. The number of Lyapunov exponents for an attractor in *d*-dimensional phase space is equal to the number of dimensions for that phase space.

A chaotic attractor has negative exponents associated with a contraction in phase space and at least one positive maximal Lyapunov exponent that measures the level of exponential divergence of small volumes in phase space, which accounts for the phenomenon of sensitive dependence upon initial conditions and, in the context of stochastic chaos, of noise amplification, being a key signature of chaos.

A convergence of Eckmann *et al.*’s estimated Lyapunov spectrum for the phase space reconstructed trajectory, with at least one positive Lyapunov exponent is, thus, evidence of chaos [20].

Using the estimated Lyapunov spectrum, we also calculate the Kaplan-Yorke dimension [21], which is a fractal dimension that uses the Lyapunov spectrum. The dimension calculation is based on the following formula:

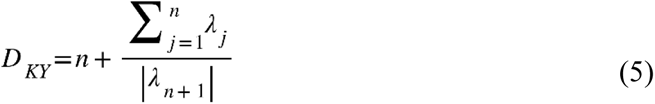

In equation (5), the *λ_j_* are the Lyapunov exponents and *n* is the maximum number of exponents, arranged in decreasing order, that can be added before the sum becomes negative. Another fractal dimension that we calculate is the box-counting dimension which provides for an additional metric on the fractal geometry of an attractor [22].

We can expect the Kaplan-Yorke dimension and the box-counting dimension to lead to values close to each other. In the context of stochastic chaos, when the stochastic component has a complex dynamics it may contribute to the fractal structure of the attractor affecting the fractal dimension, a phenomenon that we will evaluate for the case under study.

Besides the above analyses, we also perform spectral analysis and analyze the power spectrum for the original epidemiological time series and the reconstructed attractor for each phase space coordinate.

This analysis is important to distinguish the type of chaos that one may be dealing with, namely, chaotic dynamics can have white noise spectra, also called white chaos, but can also have other types of power spectrum signatures including power law decaying spectra, that is, power spectra that are close to power law noise or 1/f noise [11], this type of chaos is also known as color chaos [5,12].

The power law scaling associated with long-range dependence tends to counter the exponential divergence associated with positive Lyapunov exponents, in this way, chaotic dynamics with long-range dependence and power law scaling in the power spectrum, may have positive low Lyapunov exponents, thus, weak chaos is expected.

The emergence of finite-dimensional attractors in complex systems corresponds to a phenomenon of emergence of a finite number of collective degrees of freedom that end up driving the system’s dynamics as order parameters in Haken’s sense [23].

These collective degrees of freedom lead to the main Euclidean dimensions for the emergent chaotic attractor, in this context, if the system’s dynamics is characterized by a power law scaling in the power spectrum, despite being chaotic tends to be weakly chaotic and have a stochastic component that is, stochastic chaos is expected.

In the case of COVID-19’s main epidemiological series, the type of chaos that seems to emerge is characterized by low-dimensional stochastic chaotic attractors with power law scaling and low values of the largest Lyapunov exponent, near 0.001 for the regional data [5].

We will see that a similar phenomenon occurs for the influenza data under analysis in the present work, with the estimated largest Lyapunov exponent also being equal to 0.001 in a three decimal places’ approximation, with power law scaling in the power spectrum.

This scaling in epidemiological series and weakly chaotic dynamics are linked to contagion processes that produce strong temporal correlations, which lead to the simultaneous occurrence of power law scaling in the power spectrum and weakly chaotic dynamics, these are actually two linked statistical markers with an epidemiological basis [5].

Besides the above analyses, we also use topological data analysis methods, which, in our case, will involve recurrence analysis and *k*-nearest neighbors’ graph analysis.

Recurrence analysis takes advantage of the Euclidean topology of the reconstructed phase space, in this case, we can calculate the distance matrix which records the Euclidean distances between each phase point, formally, the matrix entries are given by the Euclidean norm:

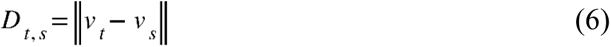

From this matrix, a colored recurrence plot can be obtained with the color range reflecting the distances. Given the distance, a recurrence event for a given radius can also be calculated by registering the value 1 if the Euclidean distance is at most equal to that radius and 0 if it surpasses that radius, this leads to the radius-dependent recurrence matrix with entries [5,18]:

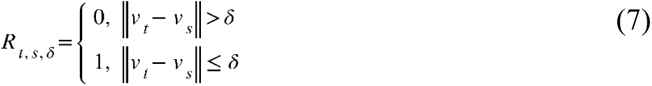

This is a square symmetric matrix that registers the value 1 when two phase points do not differ by more than the radius, and 0 otherwise. While the open neighborhood could be used, the closed neighborhood definition for the recurrence event has the advantage of allowing us to identify fully periodic dynamics so that, in the case of a periodic dynamics, if the radius is equal to zero, all diagonals parallel to the main diagonal that differ from each other by the period in question will have the value of 1 in each matrix entry, otherwise the value will be 0.

In the nonperiodic case, we do not get these evenly spaced full diagonals; when the radius is zero, the nonperiodicity leads to a value of 0 in the recurrence matrix for every point, since no phase point repeats itself.

In the analysis of the recurrence patterns, we calculate three statistics for different radii in order to better characterize the resulting dynamics [5]. The first statistic is the recurrence probability that measures the probability that a randomly selected diagonal below the main diagonal of the recurrence matrix has recurrence points. This probability can be calculated by dividing the total number of diagonals below the main diagonal with recurrence points by the total number of diagonals below the main diagonal. Since the recurrence matrix is symmetric, only the diagonals below the main diagonal are counted; alternatively, one could perform the same calculation using the diagonals above the main diagonal.

The second statistic that we will use is the average recurrence strength, which corresponds to the sum of the number of points that fall within a distance no greater than the radius in each diagonal below the main diagonal, divided by the total number of diagonals below the main diagonal with recurrence. This measure evaluates how strong on average the recurrence is.

The third statistic that we will use is the conditional 100% recurrence probability: this is the probability that a randomly chosen diagonal line with recurrence has 100% recurrence, for the radius chosen, this metric allows us to identify possible resilient periodic or quasiperiodic signatures.

If all lines with recurrence had 100% recurrence for the radius chosen, then this number would be equal to 1, the lower this statistic is, that is, the closer to zero it is, the more interrupted the diagonals are, this metric should be read in conjunction with the previous two [5].

The second type of topological data analysis method uses the *k*-nearest neighbors’ graph analysis (KNN graph) in order to characterize the attractor’s topological properties. As in the recurrence analysis, we calculate the main statistics for different neighborhood sizes, in the case of the KNN graph analysis we build a KNN graph for different *k* and analyze the graph’s main topological properties.

As an increase in the radius in recurrence analysis leads to a picture of greater neighborhood size, the increase in *k* also corresponds to an increase in the neighborhood size. For each value of *k*, each phase point is linked to its *k* nearest neighbors in phase space, which provides for a graph-based representation of the attractor’s neighborhood configuration.

For this graph, that we denote by *G*, given the set of alternative degree values *S* and relative frequencies *f*(*s*) for each degree value we calculate the relative degree entropy [5]:

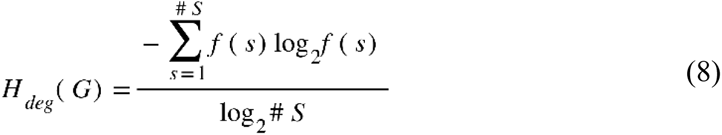

In the above equation, one is dividing the Shannon entropy for the graph’s degree distribution by the maximum entropy that one would obtain for a random graph distribution, in this way, we get a relative entropy, between 0 and 1, where the value 1 would correspond to a maximum entropy degree distribution.

In the case of a chaotic attractor, the relative entropy can actually be low, especially for weak chaos, and rise with the number of nearest neighbors used in the calculation of the graph.

This is expected since, as *k* is increased, new recurrences start to appear leading to a more random graph distribution due to the fact that chaotic dynamics can lead each orbit close to each other. A stochastic component can also sometimes introduce proximity between phase points due to noise.

The other graph entropy measure that we calculate is the graph’s Kolmogorov-Sinai (K-S) entropy, which provides for a graph complexity measure.

Indeed, the K-S entropy is an information measure on the sequence of nodes for a Markov process on a network, in this case, for a KNN graph it provides an information measure for a Markov process with a transition matrix extracted from the *k*-nearest neighbors.

For an unweighted graph, which is the case that we are dealing with, the entropy coincides with the logarithm of the dominant eigenvalue of the transition matrix, expressing it in bits leads to the information measure [5]:

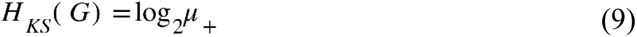

Where the argument in the logarithm corresponds to the referred dominant eigenvalue.

The third metric that we calculate is the Wiener index for the KNN graph, which corresponds to the sum of the lengths of the shortest paths between all pairs of phase points linked in the KNN graph.

In the case of chaotic dynamics, we expect the two entropy measures to rise with *k* and the Wiener index to actually drop, since as we increase the neighborhood size recurrences start to appear and the paths start to become shorter. The Wiener index tends to have higher values in more chaotic attractors and lower values in weaker chaotic attractors.

While the above analysis provides for a topological characterization of the system’s phase space dynamics, including a possible chaotic attractor, it is insufficient for our main objective in terms of analysis, which is to address the dynamics for the epidemiological target variable, including both possible chaotic dynamics and stochastic dynamics, that is, we wish to capture all the pattern and characterize it down to independent and identically distributed noise (IID noise), this is where phase space decomposition is used in conjunction with the above topological data analysis methods [13].

Formally, the time series can be effectively divided into two components, the agent’s predictions and the residuals component, using equation (4) the values for the series decompose as:

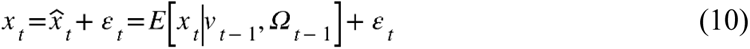

The delay embedding of the series is, therefore, embedding both components the deterministic component captured by the adaptive topological agent and the residuals, therefore, such as the series decomposes exactly in the predictions plus residuals, the dynamical phase point does so as well so that, replacing in equation (2), we get the phase space decomposition [13]:

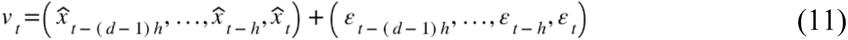

The most elementary case is one in which the residuals are IID noise, however, in complex systems’ dynamics this is not always the case, namely, the residuals may be described by a stochastic process which may exhibit turbulent and even nonlinear dynamics, there is also the possibility of feedback from a chaotic signal to the stochastic component in which case, the process becomes more complex and also has a deterministic element to it. The possibility of multiple chaotic attractors can also occur when dealing with such residuals [13].

In this way, we need to address the phase space decomposition and test the residuals component for chaos and research its structure. The modeling of the residuals is an important step in order to address stochastic chaotic dynamics.

The topological analysis of the impact of the residuals in the dynamics is also important; in this sense, the analysis of the filtered dynamics obtained from the adaptive topological agent’s predictions versus the original attractor is a necessary step.

While most studies have focused on finding and characterizing a fractal chaotic attractor using a noise filter and do not address the noise, filtering it out of the analysis, in the context of stochastic chaos, the research of the stochastic process down to an IID component is important for a full characterization of the system’s complex nonlinear dynamics including the chaotic component that was filtered, in our case, using the *k*-nearest neighbors’ structure, and the stochastic component which is left out and that can have itself a complex pattern that needs to be addressed.

The testing for IID noise can be done by the Brock, Dechert and Scheinkman (BDS) test which tests the null hypothesis that the series is IID [24], another statistical test is Engle’s test that tests for a type of nonlinear dependence that is called autoregressive conditional heteroskedasticity (ARCH) for which the noise variance is nonstationary in an autoregressive way [24].

Engle’s test addresses quadratic autocorrelation in a series’ residuals, the null hypothesis is that no such autocorrelation exists, the alternative is that there is autocorrelation and therefore ARCH-type nonlinearity, there are two versions of the test one that uses Lagrange multipliers and another that uses the F statistic. We will use here both the BDS test and the two versions of Engle’s test in conjunction.

Capturing the IID noise and studying its probability distribution further allows us to work on the probabilistic study of a stochastic chaotic dynamics, which in the epidemiological context is an important factor that we illustrate in the present work.

We apply these methods to the FluNet dataset by the World Health Organization processed by Our World In Data in the indicator *weekly share of positive tests – all types of surveillance*, which involves calculating, on a weekly basis, the share of positive tests to any influenza strain for different regions and countries, the region we will be researching in the present work is the Northern Hemisphere. The period under analysis is from 2009-01-05 to 2025-03-31.

Applying the above methodology, we are able to decompose the above series into its various components down to the IID noise factor and the corresponding probability distribution. This allows us to perform a probabilistic analysis of the series’ major patterns and illustrate how the methodology can be employed for risk analysis in epidemiological studies focusing on target epidemiological indicators.

## 3. Main Results

In figure 1, we show the weekly share of positive tests to influenza in the Northern Hemisphere from 2009-01-05 to 2025-03-31, which was the last data point available at the time of the analysis. As can be seen from the graph, there are multiple spikes associated with outbreaks, interspersed with low detection periods, which is characteristic of an intermittent outbreak pattern.

**Figure 1:**
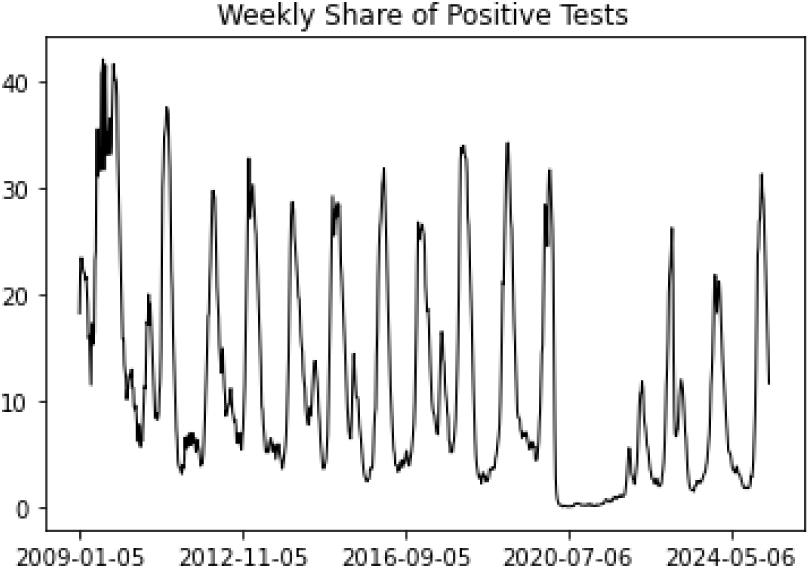
Weekly share of positive tests to influenza for the Northern Hemisphere in percentage. Source: FluNet by the World Health Organization (2023) – processed by Our World in Data from 2009-01-05 to 2025-03-31.

Some underestimation of the actual number of infections is expected, however, since we are dealing with an aggregate epidemiological indicator, specific testing differences are averaged out leading to an overall picture that is strongly linked to epidemiological processes and contagion dynamics, so that the aggregate indicator for the Northern Hemisphere and all influenza types provides for a correlated indicator with actual underlying contagion dynamics and corresponding epidemiological processes for influenza.

Noticeable in the graph is a drop during the COVID-19 lockdown; we will find that this drop is actually captured at the joint level of both the chaotic process and dynamical standard deviation of the nonlinear stochastic process itself.

In figure 2, we show the power spectrum for the series. We find that there is a power law decay in the power spectrum with a fitted slope of -2.5158, for an *R*^2^ score of 69.32% and a *p*-value of 1.7191e-107.

**Figure 2:**
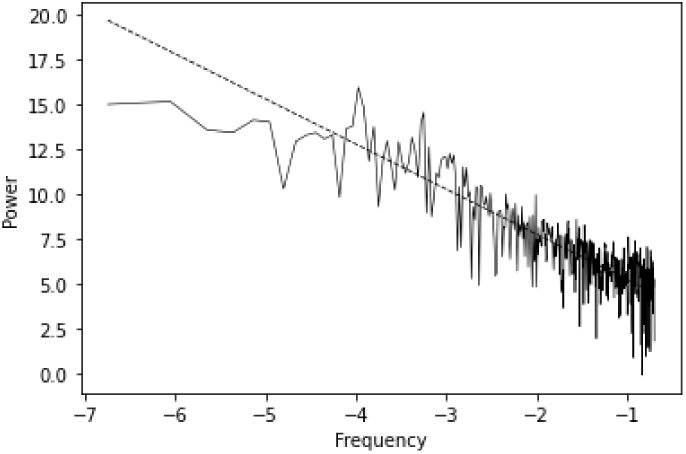
Power spectrum for figure 1 series along with the fitted slope.

These results are indicative of a power law noise-like spectrum corresponding to black noise, which is a type of dynamics that shows strong persistence and long fractal memory that occurs in different risk dynamics, including natural disasters [26].

Black noise spectra also occur in the epidemiological context [5], which is to be expected and can be linked to the epidemiological outbreak dynamics, with periods of low contagion followed by periods of accelerating contagion, leading to a spike of detections followed by a subsequent reduction in cases, producing an intermittent dynamics with persistence and long memory. We will return to this point further on, in regard to the underlying chaotic attractor dynamics itself.

Proceeding now to the embedding parameters’ selection, and since we will be using the KNN graphs for attractor topological analysis, as explained in the previous section, we use an adaptive topological learner equipped with a *k-*nearest neighbors’ learning unit. Employing a sliding learning window of 10 weeks, and *k* equal to 5 nearest neighbors, which is half of the sliding learning window size, allows us to capture enough local topological pattern without adding excess pattern in terms of neighborhood size. The weights used are based on the Euclidean distance, which is also used for the KNN graphs and the recurrence analysis. The learning algorithm used is the *KD Tree*.

The setting of the parameter to *k* equal to 5 nearest neighbors captures a five-week pattern, which encompasses a monthly dynamics and the transition between two months. The 10 weeks’ sliding learning window allows for one to capture bimonthly patterns and a transition week along with the patterns within that week, which is effective when dealing with the exploitation of topological patterns in the context of intermittent chaotic dynamics, which, as we will see, characterizes the influenza dynamics.

The number of tested dimensions were from 1 to 20 and the lags also from 1 to 20. The optimal embedding selected, using the *R*^2^ score as a performance metric, was found for a two-dimensional embedding and a lag of 11 weeks (table 1), which is actually consistent with the learning window size for pattern capturing. It is important to stress that the topological adaptive agent is rebooted for each tested embedding so that no pretrained agent is used in multiple tests.

**Table 1:** Optimal embedding parameters obtained for the adaptive topological agent, corresponding prediction performance metrics, estimated Lyapunov exponents and Kaplan-Yorke dimension.

|  |  |
| --- | --- |
| Lag | 11 |
| Dimension | 2 |
| $R^2$ Score | 78.33% |
| Explained Variance | 78.34% |
| Correlation | 0.8902 |
| RMSE | 4.7277 |
| RMSE/Amplitude | 11.23% |
| L1 | 7.1086E-04 |
| L2 | -8.2785E-04 |
| DKY | 1.8587 |

Before we analyze table 1’s results, we first look at the pattern of performance for the optimal lag with different embedding dimensions, to understand how the exploitable topological information linking the attractor’s previous phase point to the next value of the series behaves with the dimension, this is shown in figure 3.

**Figure 3:**
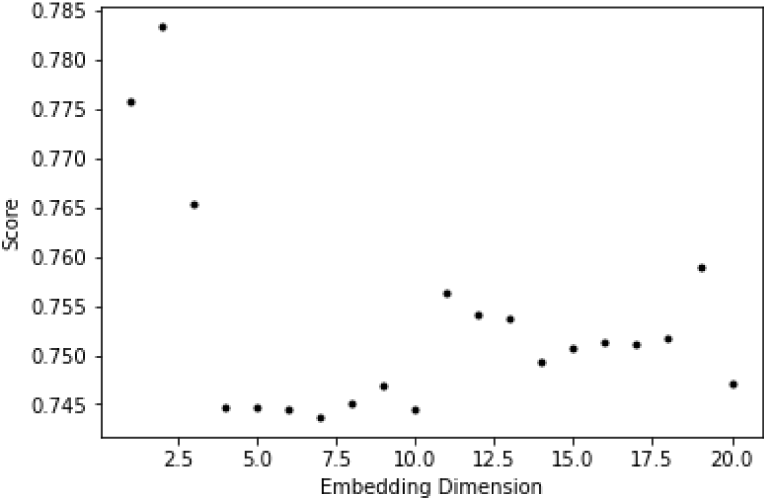
Adaptive topological agent’s *R*^2^ score as a function of the embedding dimension for the lag 11.

As can be seen, from figure 3, the best performance is obtained for low dimensions 1 to 3, with 2 being the dimension where the performance is highest, that is, the dimension for which there is more exploitable topological information linking the series to the reconstructed phase space trajectory.

As the embedding dimensions are increased, the score for the adaptive topological agent is lower which means that there is exploitable topological information lost by increasing the embedding dimension beyond the threshold of 3. However, all of the dimensions lead to an *R*^2^ score larger than 0.7 which indicates a more than 70% ability to predict the target series from the embedded trajectory, indicating a strong topological structure for the optimal lag.

In figure 4, we compare the different tested lags in terms of optimal *R*^2^ scores and corresponding embedding dimensions, with the labels at each point in figure 4’s graph showing the corresponding optimal dimension. As can be seen in figure 4, the prediction scores are all high, above 77%; however, the highest values are obtained for dimension 2 and for the lags between 7 and 13. Also, dimension 2 is the dominant dimension, appearing more times as the optimal dimension.

**Figure 4:**
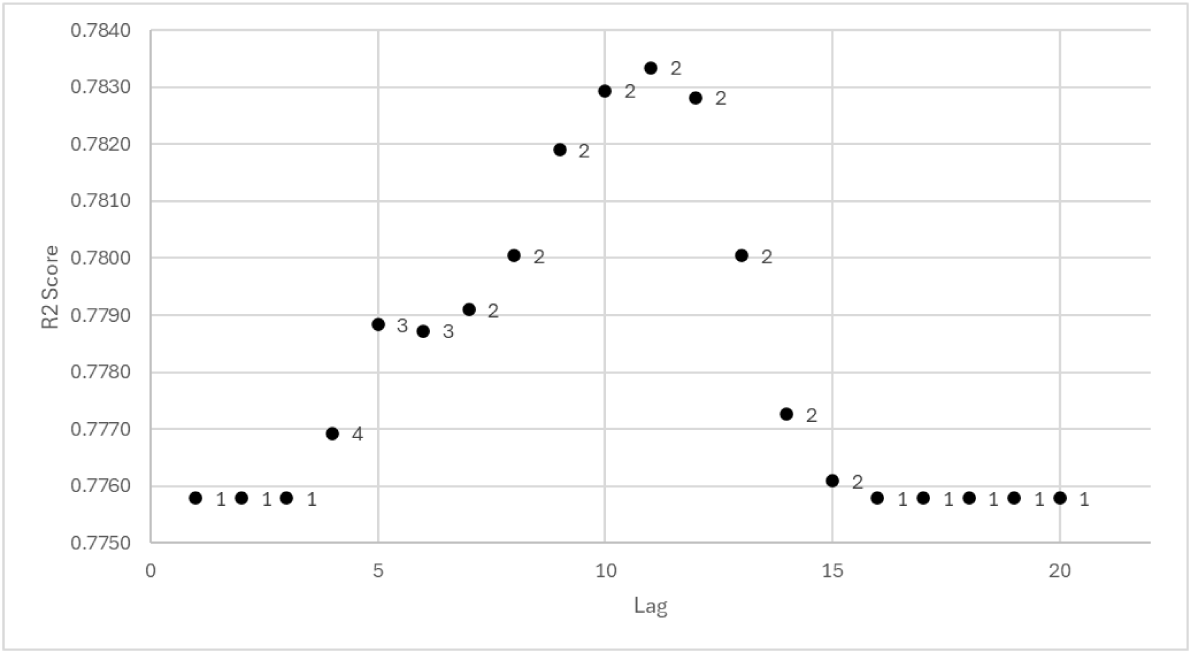
Adaptive topological agent’s highest *R*^2^ score as a function of the tested lag, the labels are the corresponding optimal dimensions for which the score was obtained.

The other embedding dimension appearing more frequently after 2, is 1 dimension, which occurs for low lags, 1 to 3, and high lags, 16 to 20. The dominance of low dimensions for the tested lags is strongly indicative of an emergent dynamics characterized by an underlying low-dimensional attractor.

Indeed, for all the tested lags, the optimal dimensions all range between 1 and 4, with 2 being the dominant dimension in terms of appearing more often with the highest *R*^2^ score, and the one for which the best performance is obtained.

The highest prediction score is obtained for lag 11 with an *R*^2^ of 78.33%, an explained variance of 78.34%, a correlation of 0.8902 between the adaptive topological agent’s predictions and the target epidemiological variable, an RMSE of 4.7277 which corresponds to a 11.23% error relative to the total amplitude of the series, as shown in table 1.

The high values in the topological agent’s performance metrics for the reconstructed attractor indicate that there is a strong exploitable topological pattern linking the reconstructed attractor dynamics to the weekly share of positive tests to influenza.

In figure 5, we show the observed series in blue and the predicted series in orange, as can be seen, from figure 5, the adaptive topological agent is able to capture well the overall dynamics, which indicates that there is a strong topological information that can be exploited for predicting the target series from the embedding results, this is confirmed by the above analysis of table 1’s prediction performance metrics.

**Figure 5:**
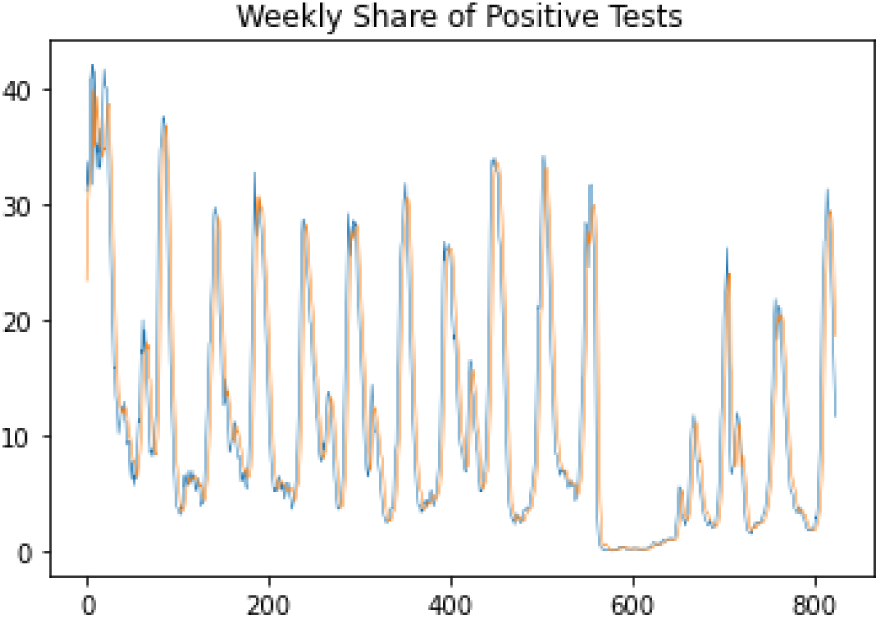
Adaptive topological agent’s predictions (orange) and observed values (blue).

Analyzing the reconstructed attractor we also find that the Lyapunov spectrum converges (figure 6) with a positive Lyapunov exponent of around 7.1086E-04 and a negative exponent of around -8.2785E-04, leading to a Kaplan-Yorke dimension of 1.8587.

**Figure 6:**
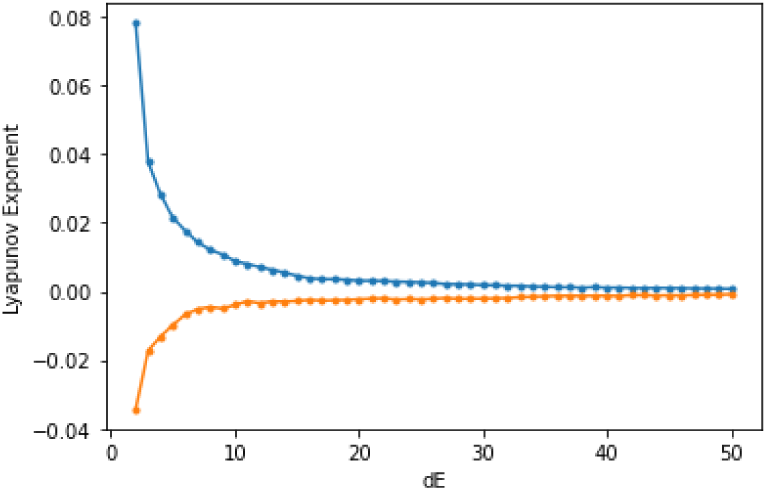
Estimation of the Lyapunov spectrum for the reconstructed attractor, using the matrix dimension as 2 and the lag 11.

In this way, the evidence is favorable to a strange chaotic attractor with a fractal dimension between 1 and 2 but closer to 2. As we will see, the Kaplan-Yorke dimension is in good agreement with the box-counting dimension.

These results are consistent with a two-dimensional map rather than a flow, that is, a discrete nonlinear dynamical system, since, for flows, the minimum number of phase space dimensions necessary for chaos is three.

The low value of the largest Lyapunov exponent indicates that the dynamics is characterized by weak chaos, if we estimate the same exponent for the embedded adaptive topological agent’s predictions using that embedding as a noise-filtered phase space trajectory, as explained in the previous section, we also find a convergence, as can be seen in figure 7, however, the value for the largest exponent is slightly higher, in this case, the estimated positive exponent is 9.1216E-04, which is closer to 0.001, and the estimated negative exponent is -9.3283E-04, the values of the two exponents are very close to each other, which indicates that we have a weakly dissipative chaotic dynamics with a Kaplan-Yorke fractal dimension of 1.9778 which is very near the dimensionality of the two-dimensional Euclidean phase plane in which the attractor “lives”.

**Figure 7:**
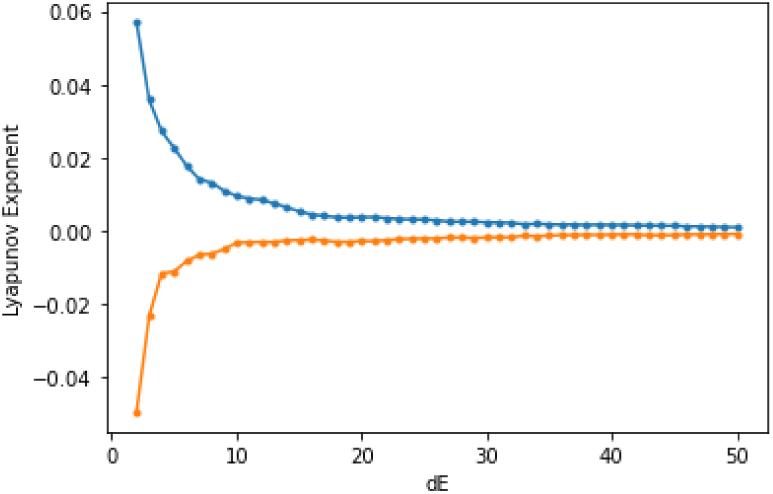
Estimation of the Lyapunov spectrum for the embedding of the adaptive topological agent’s predictions, using the matrix dimension as 2 and the lag 11.

The weak chaos is expected, given the black noise-like power spectrum of the main series. The evidence, as we will see through the application of the topological data analysis methods, is that we have a dynamics that is very near the onset of chaos with a marginally stable fixed point, which explains both the power law scaling in the series and is consistent with the overall dynamics.

Epidemiologically, this is linked to the outbreak process by which there are periods of low contagion and low detections, and then these periods are interrupted by accelerating contagion waves leading to spikes in detection and an overall intermittent chaotic dynamics. In this way, we expect, in the recurrence analysis, fixed-point signatures to show up, indicating that the dynamics is very near the onset of chaos, with a transition between a fixed point and a chaotic dynamics. We will return to this point further on.

In figure 8, we show the attractor reconstruction resulting from the embedding of the original series and of the adaptive topological agent’s predictions. As can be seen in the figure, the attractor shows a conic structure with the vertex near the origin, where there is a clustering of points.

**Figure 8:**
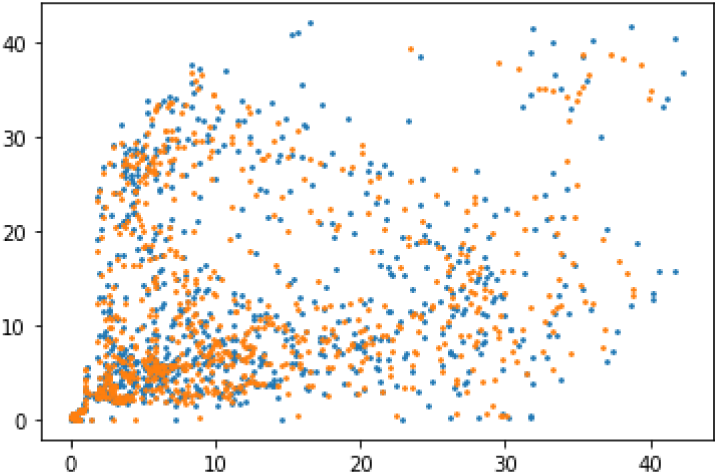
Phase space embedding of the original series (blue) and of the adaptive topological agent’s predictions (orange).

The main attractor structure further reinforces the type of dynamics captured by the adaptive topological agent; namely, we have a clustering around the origin, which corresponds to the region of low-contagion, and then the attractor forms a cone-shaped structure, which is consistent with a bottleneck effect and intermittency with long-range dynamics.

This long-range power law scaling dynamics characterized by a black noise-like spectrum is also present for each phase space dimension, both for the original attractor and for the filtered attractor reconstructed from the adaptive topological agent’s predictions, which corresponds to the predicted attractor shown in figure 8 in orange.

As shown in figure 9, the power law scaling that we found for the main series’ power spectrum is present for both phase space dimensions in the original attractor and the predicted attractor obtained from the embedding of the adaptive topological agent’s predictions.

**Figure 9:**
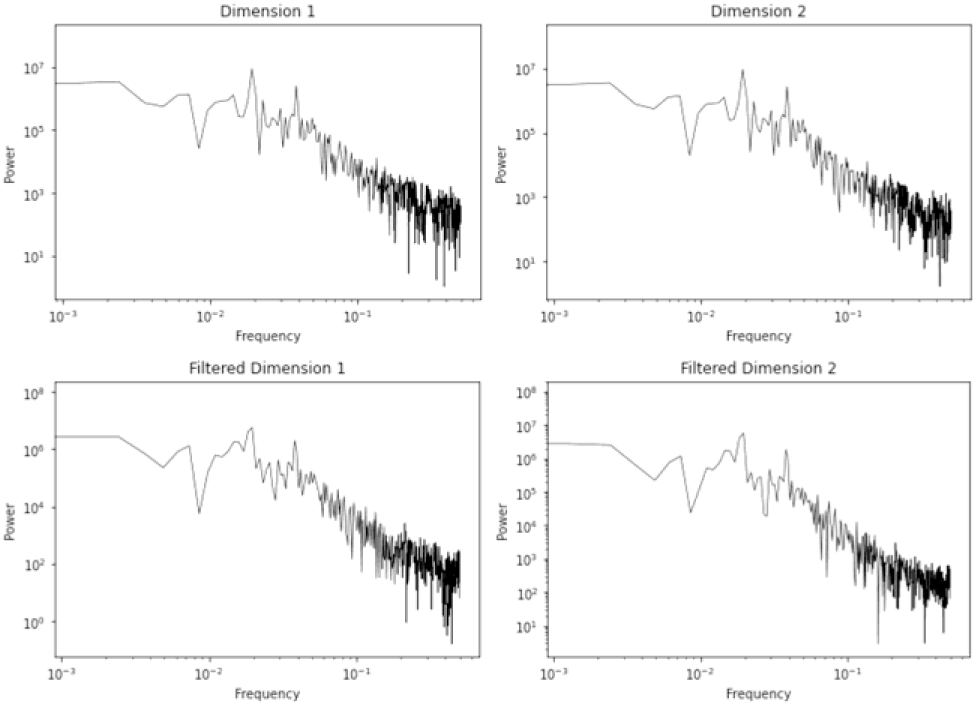
Power spectrum for the series of values for the two phase space dimensions for the original reconstructed attractor (top) and for the filtered attractor obtained from the embedded topological adaptive agent’s predictions (bottom).

**Table 2:** Estimated power law decay exponents for the 1/*f^B^* spectrum from the different estimated spectra and respective *p*-values and regression *R*^2^.

| | Exponent | p-value | $R^2$ |
| --- | --- | --- | --- |
| Main series | -2.5158 | 0.0000 | 69.32% |
| Dimension 1 | -2.5525 | 0.0000 | 73.26% |
| Dimension 2 | -2.5848 | 0.0000 | 75.32% |
| Filtered Dimension 1 | -3.1226 | 0.0000 | 77.48% |
| Filtered Dimension 2 | -2.6127 | 0.0000 | 77.56% |

The predicted attractor actually has higher values for the power law exponents than both the original series and the original reconstructed attractor, which indicates that the residuals’ component actually reduces the long-range persistence, indicating that this persistence is mainly linked to the chaotic attractor itself, showing a very strong persistence with exponents higher than 2.

Therefore, the spectral analysis results are consistent with a type of black chaos, that is, a type of color chaos characterized by black noise-like spectrum, that is, chaos with strong persistence and long-range dependence, characterized by fractal memory with a power law decay in the power spectrum with a power law exponent greater than 2 for all phase space dimensions.

The black noise-like signature in the original series thus seems to come from the underlying chaotic attractor, given the fact that the filtered dynamics obtained from the predicted attractor has black noise-like signatures for both phase space dimensions.

There is a twofold consequence from black chaos. On the one hand, the long-range dependence compensates for the exponential divergence associated with the chaotic dynamics, but it also means that amplified deviations have long-range consequences. The black chaos dynamics is consistent with other epidemiological series that show evidence of a similar underlying dynamics [5].

As stated, the black chaos phenomenon is associated with a specific epidemiological pattern that involves periods of low contagion followed by periods where contagion starts to accelerate, generating epidemiological waves and producing an intermittent pattern. This epidemiological dynamics leaves a mark both at the level of the chaotic component and at the level of the stochastic component for the residuals’ process that, as we will see, shows evidence of turbulence and also intermittency in the nonlinear dynamical variance.

The testing process for influenza also plays a role here, in the sense that lower contagion phases can lead to underreported cases, which may lead to longer lower detection rates and thus push the statistics to a dynamics that is nearer the onset of chaos, with a transition between a fixed point of very low detection and chaos, which translates, in this case, to longer-lasting low detection periods followed by accelerating detection rates. This process further increases the fractal persistence of the dynamics and may also explain the weakly dissipative dynamics with a Lyapunov spectrum sum that is nearer zero.

Like the Kaplan-Yorke dimension, the box-counting dimension is between 1 and 2, which is consistent with a fractal attractor, in the case of the observed embedded series the estimated dimension is 1.7891 with an associated *p-value* of 0.0, while for the embedded predicted series it is 1.8032, also with an associated *p-value* of 0.0. The values are close to those obtained for the Kaplan-Yorke dimension, though slightly lower.

In figure 10, we show the estimation of both box-counting dimensions with the linear fit on the log-log graph estimated on the linear region. There is a breakdown at the later part of the graph which tends to occur in nature due to finite scaling of data [22,26].

**Figure 10:**
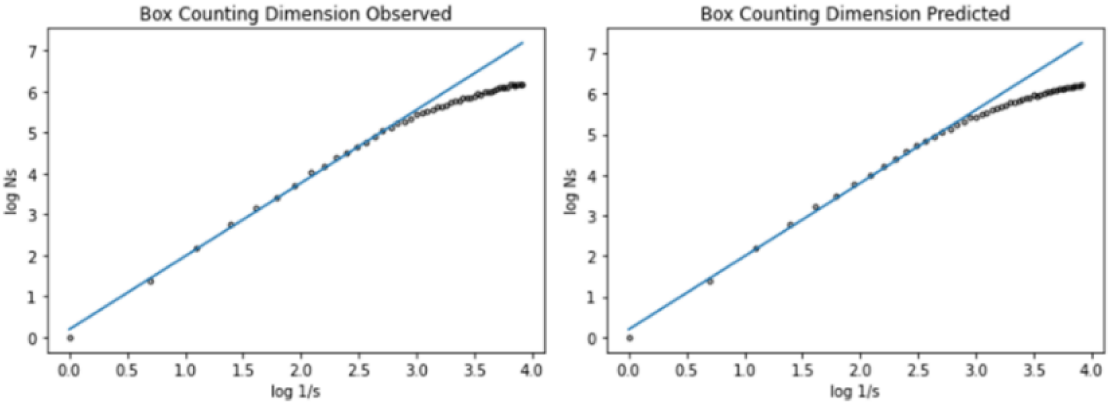
Box-counting dimension for the original attractor (left) and for the attractor obtained from the embedded predicted series (right), 50 boxes were used, the regression was performed on the linear scaling region.

The predicted attractor, obtained from the embedding of the adaptive topological agent’s predictions, exhibits a higher fractal dimension than the original reconstructed attractor, however, the effect is not too large, in the case of the box-counting dimension both the original attractor and the predicted attractor have the same dimension value in a two-decimal place approximation.

This last result indicates that while the residuals’ component, as per equations (10) and (11), has some effect on the fractal geometry of the stochastic chaotic attractor, that effect is not significant; the topological metrics are also not significantly affected by the residuals’ component.

Indeed, the degree entropy and the K-S entropy for the KNN graphs of the original attractor and the predicted attractor, which effectively captures the chaotic component, have a similar profile with values close to each other, as shown in Figure 11.

**Figure 11:**
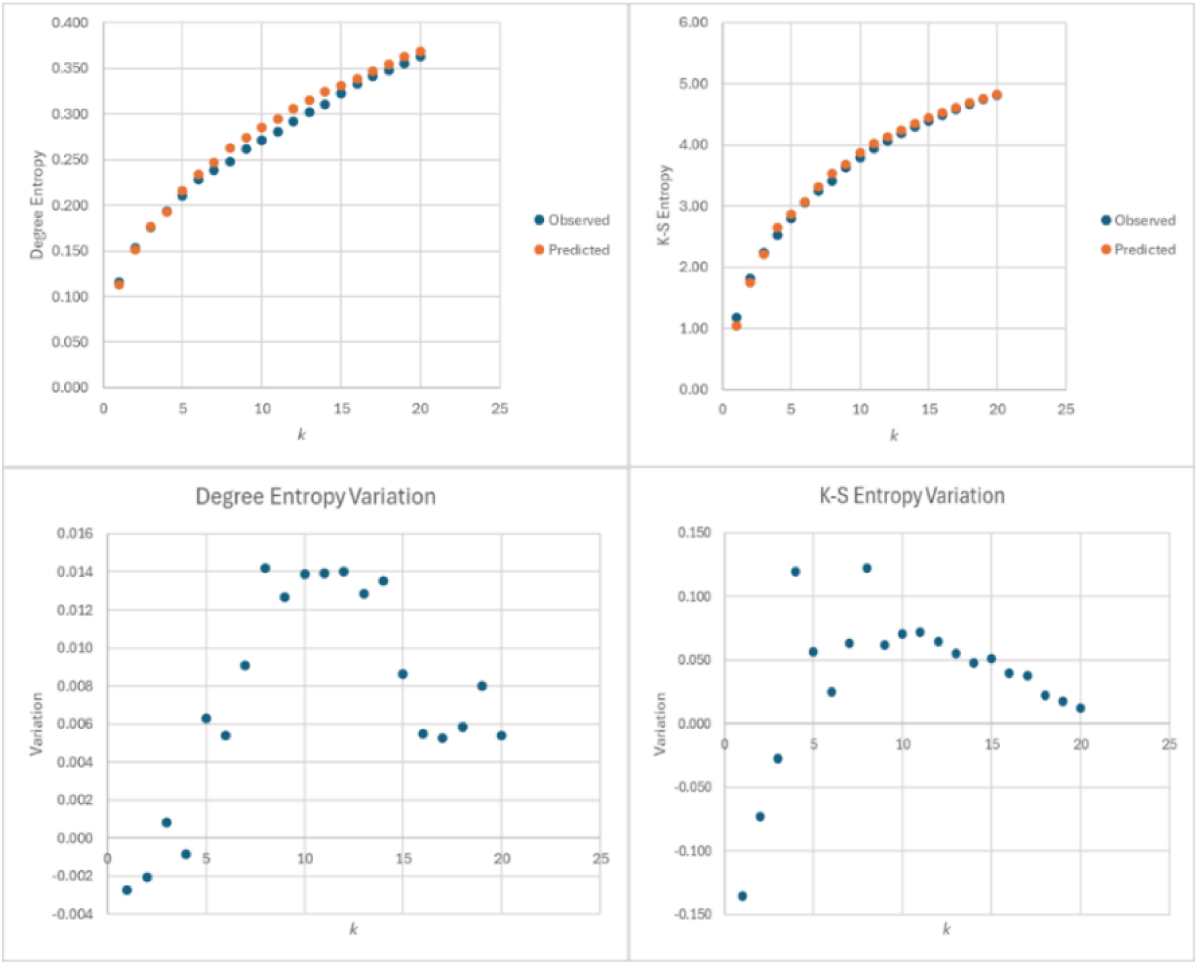
Entropy measures of the KNN graphs for the observed attractor and the predicted attractor (top) and variation from the predicted attractor to the observed attractor (bottom), for *k* increasing from 1 to 20 in steps of 1.

The degree entropy is low and tends to increase with the number of nearest neighbors as expected since more connections appear as the value *k* is increased, which is equivalent to increasing the neighborhood size.

As results from Figure 11 (bottom), for *k* equal to 1, 2 and 4, the degree entropy of the predicted attractor is lower than that of the original attractor, the same happens for the K-S entropy for *k* equal to 1, 2 and 3.

Which indicates that, for low neighborhood size, the predicted attractor tends to have lower entropy values than the original attractor. In this way, for low neighborhood size, the residuals’ component in phase space leads to an increase in entropy of the KNN graphs, which indicates a more complex neighborhood structure; however, the differences are small, which shows that the residuals do not significantly affect the main topological structure of the underlying attractor.

For intermediate values of *k*, the entropy differences tend to be higher, with the predicted attractor having higher entropy values than the original attractor, but as *k* is increased further these differences tend to become smaller. Overall the maximum absolute difference in degree entropy is 0.0142 and the minimum absolute difference is 0.0008, which shows that the impact of the residuals in the overall degree entropy is low.

For the K-S entropy, the differences are higher, the maximum absolute difference is 0.1356 and the minimum absolute difference is 0.0124. The difference between entropy values range from 0.1226 for *k* equal to 8 and -0.1356 for *k* equal to 1, which means that, for *k* equal to 8, the K-S entropy value of the predicted attractor’s KNN graph is 0.1226 higher than that of the observed attractor, while for *k* equal to 1 the K-S entropy of the predicted attractor’s KNN graph is -0.1356 lower than that of the observed attractor. In this way, locally the residuals’ component adds to the neighborhood complexity of the graph while for higher neighborhood sizes there is a reduction in that complexity, however, the difference between the predicted and original attractor profiles is small.

For the Wiener index, we find that both attractors have a similar overall profile. However, there are some differences. For *k* below 6, the Wiener index is not defined for the original attractor’s KNN graph due to a large number of disconnected components, while for the predicted attractor it is defined for *k* equal to 4 onwards, which indicates that the residuals’ dynamics leads to more disconnected components for low neighborhood sizes, which is consistent with the impact of a stochastic dynamics.

For values of *k* between 7 and 12, however, the predicted attractor has a higher Wiener index than the original attractor, as per figure 12 (right), which indicates that the Wiener index of the predicted attractor is higher, therefore, the dynamics is more spread out, which is consistent with the higher maximum Lyapunov exponent for the predicted attractor.

**Figure 12:**
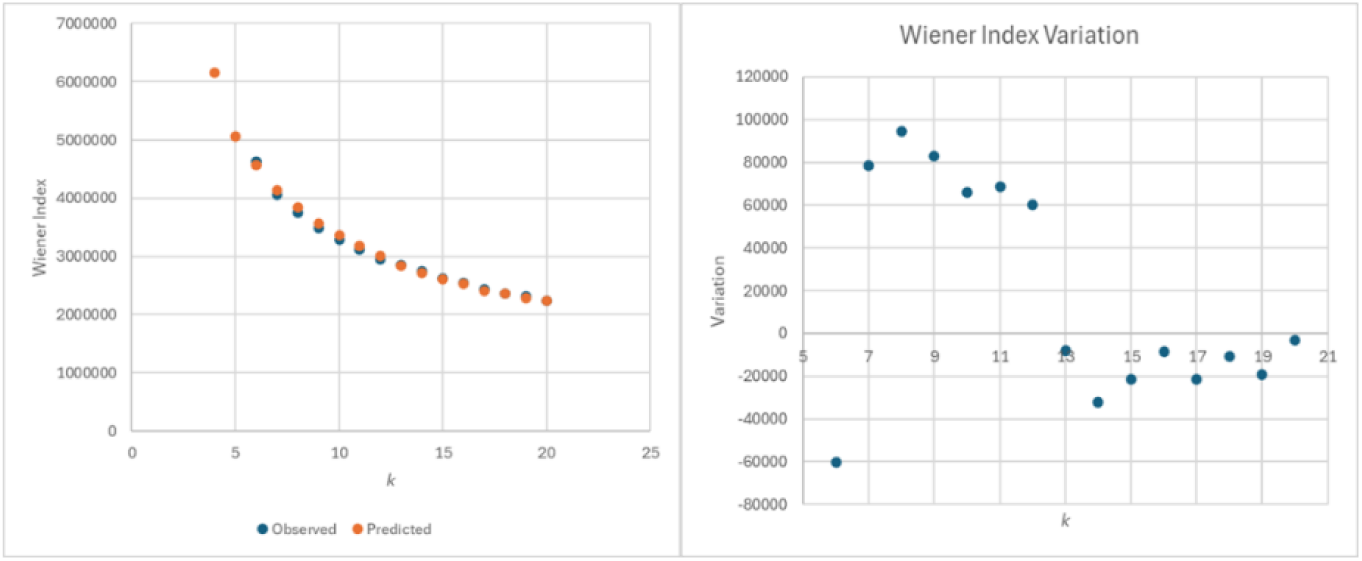
Wiener index of the KNN graphs for the observed and the predicted attractors (left) and variation from the predicted attractor to the observed attractor (right), for *k* increasing from 1 to 20 in steps of 1.

For *k* equal to 6 and for higher values of *k*, however, the relation reverses and the residuals’ component leads to an increase in the Wiener index, effectively leading to a destruction of patterns present in the original attractor.

This means that on an intermediate scale of nearest neighbors, the residuals actually add topological pattern, reducing the Wiener index for the original attractor when compared with the predicted attractor, which is consistent with the presence of possible deterministic dynamics in the residuals’ component that may add recurrence structure to the underlying attractor’s dynamics in phase space and push the dynamics closer to the onset of chaos.

As can be seen in Figure 13, the recurrence structure of both the original and the predicted attractor is similar. The recurrence metrics calculated for different radii in units of standard deviation of the original series reinforce the above analysis.

**Figure 13:**
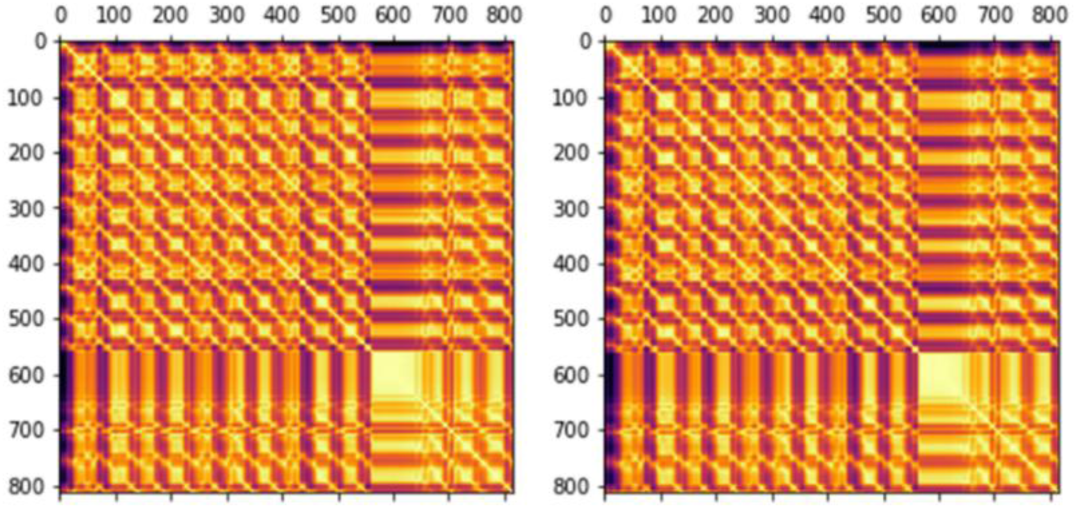
Colored recurrence plot for the observed attractor (left) and for the predicted attractor (right), Python’s Matplotlib’s reverse inferno colormap (“inferno_r”) was used so that lighter colors correspond to lower distances and darker colors to higher distances, Euclidean distance metric was used for the plot.

As per Figure 14, the recurrence probability for both the observed and predicted attractors is high, above 0.88, and it tends to increase with the radius, which is to be expected, since as the radius increases more recurrence points tend to appear.

**Figure 14:**
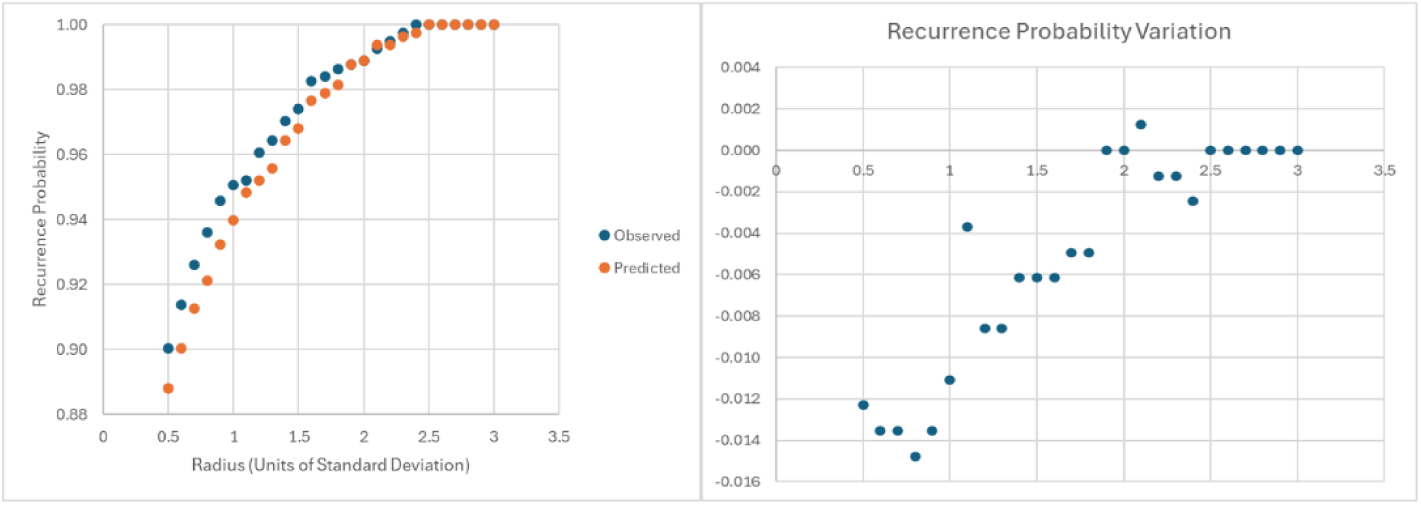
Recurrence probability as a function of the radius for the observed attractor versus predicted attractor (left) and variation of the values from the predicted attractor to the observed attractor (right), for the radius varying from 0.5 units of standard deviation to 3 units.

The high recurrence probability is consistent with the previous signatures of a chaotic dynamics near the onset of chaos. However, while similar in profile, the observed attractor has a close but higher recurrence probability for most radii than the predicted attractor and the two probabilities tend to converge with the rise in the radius, therefore, we find, again, that the residuals’ phase space component is adding a recurrence structure to the attractor’s dynamics, which indicates a non-IID noise.

While there are some differences in the recurrence probability values, the average recurrence strength of the observed and the predicted embedded series almost coincide in values, showing a linear increase with the radius, as per Figure 15.

**Figure 15:**
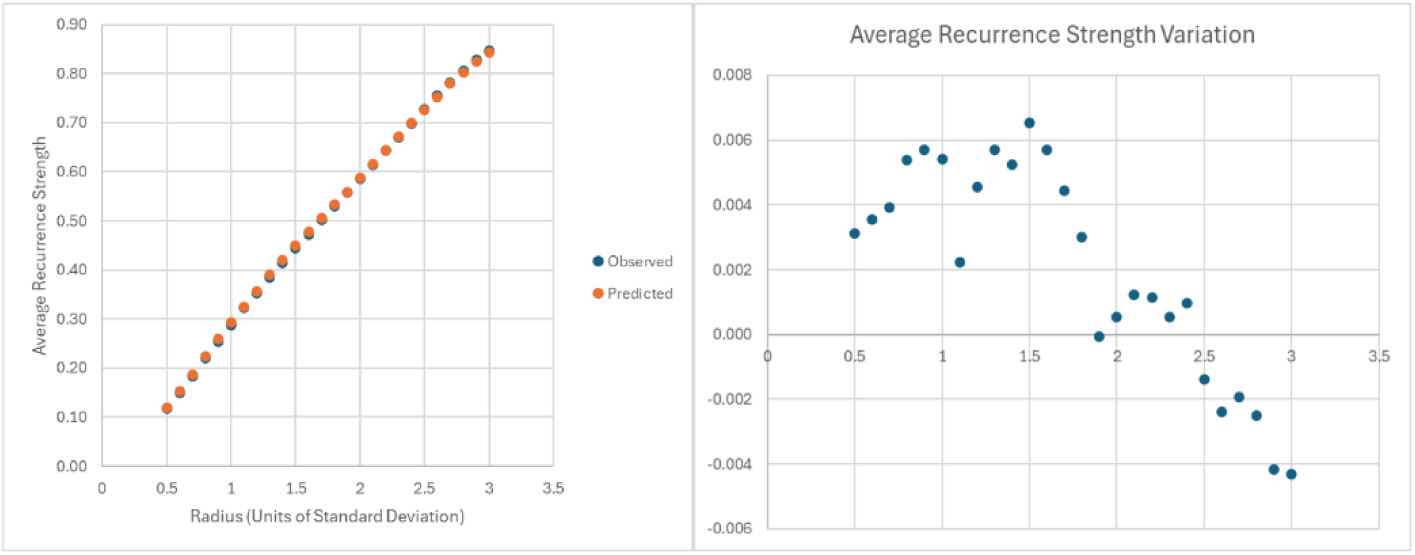
Average recurrence strength as a function of the radius for the observed attractor versus predicted attractor (left) and variation of the values from the predicted attractor to the observed attractor (right), for the radius varying from 0.5 units of standard deviation to 3.

For low radii, the average recurrence strength is small but higher than 0.1. For the lowest radius, 0.5 standard deviations, we get the lowest average recurrence strength, which is around 0.1168 for the observed attractor and 0.1199 for the predicted attractor. This means that, on average, the diagonals are filled to near 12% of recurrence points, which is consistent with interrupted diagonals typical of chaos.

For a radius of 1 standard deviation, the average recurrence strength is near 30%, which indicates a strong recurrence structure that is consistent with the markers of the onset of chaos.

For the average recurrence strength, we find, from the results shown in figure 15 (right), that, while very close to each other, for low to intermediate radii, the predicted attractor has a higher average recurrence strength than the original embedded series.

The main implication is significant: the residuals’ component actually may add localized short-range recurrence structure, but simultaneously it destroys longer range recurrences, reducing the average recurrence strength while simultaneously increasing the recurrence probability.

These results are consistent with a stochastic component that may have some structure in terms of recurrence with a complex impact on the system’s dynamics. Increasing recurrence probability and even possibly inducing predictability pockets but, simultaneously, reducing the average recurrence strength.

Now, regarding the conditional 100% recurrence probability, we find that, for low radii, this probability is zero for both the observed and predicted attractors, that is, there are no lines with 100% recurrence, which is consistent with a nonperiodic dynamics.

These lines start, however, to appear as the radius is increased following a ladder-like pattern that rises exponentially with the radius. Even though low, there is a non-null number of lines with 100% recurrence, as per figure 16.

**Figure 16:**
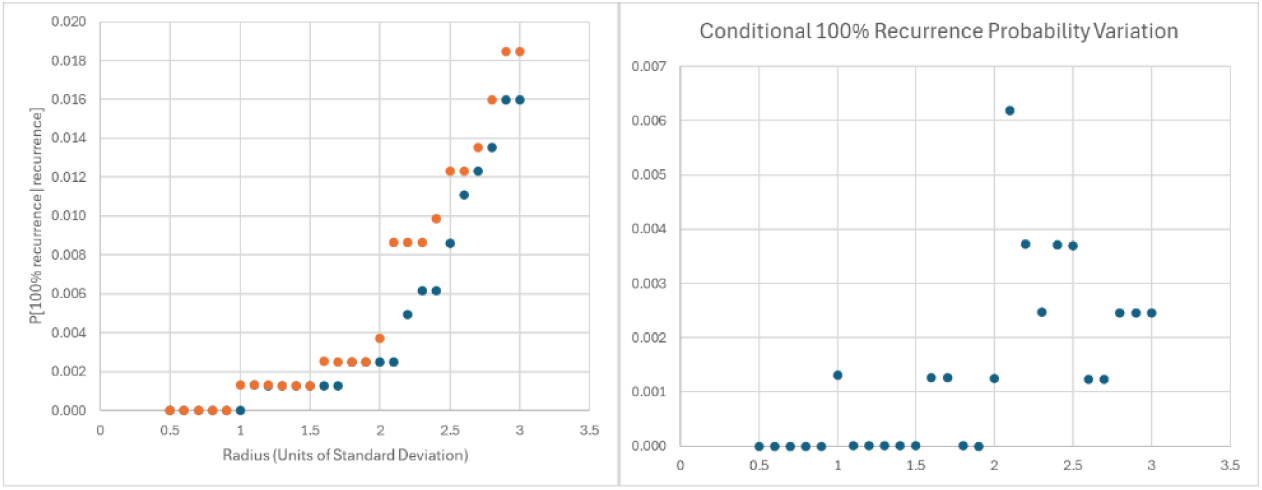
Conditional 100% recurrence probability as a function of the radius for the observed versus predicted attractor (left) and variation of the values from the predicted attractor to the observed attractor (right), for the radius varying from 0.5 units of standard deviation to 3.

In this case, these lines are associated with fixed point signatures, indeed, for a radius of 3 standard deviations, we get, for the original attractor, 13 lines with 100% recurrence, corresponding to two regions with 100% recurrence, these lines are all sequential that is they differ from each other by one iteration implying the presence of a proximity to a fixed point dynamics. For the predicted attractor these lines increase to 15 and again correspond to fixed point signatures.

As can be seen in Figure 16 (right) we find that the residuals’ component tends to reduce the probability of finding a 100% recurrence line in a random selection of lines with recurrence, though the impact is not that significant.

Having addressed the reconstructed attractor’s dynamics we now need to address the residuals’ component. From the above topological data analysis, we find that the residuals may have some structure and not be just IID noise.

Indeed, as per Figure 17, we find the presence of an intermittency and turbulence with large jumps followed by periods of lower fluctuations, typical of changing volatility dynamics, with a long low volatility period occurring during COVID-19 period, for which the influenza detections decreased. The power spectrum, as shown in figure 18, is also not a white noise spectrum.

**Figure 17:**
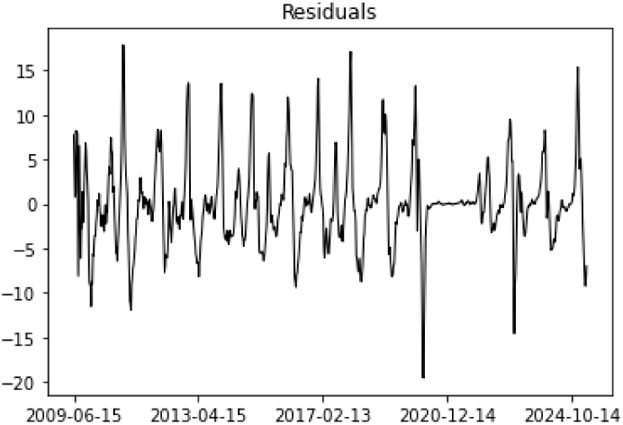
Residuals’ series from the adaptive topological agent’s predictions.

**Figure 18:**
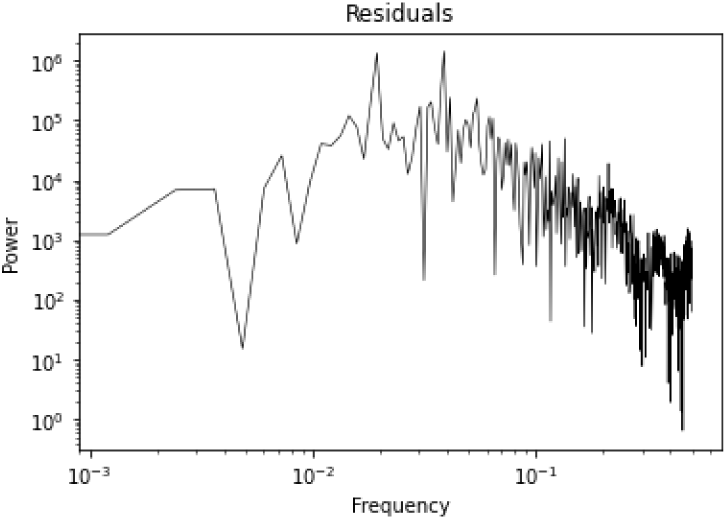
Residuals’ series power spectrum.

As per Figure 18, the power spectrum for the residuals rises with two significant spikes in the middle frequency region and then a power law decay, which shares a common feature with that of the original series.

Considering the phase space decomposition in the predicted and residuals’ component, as per equation (11), extracting the residuals’ component we get a phase space shape for that component’s dynamics with a winged structure which is associated with the changing volatility dynamics, as shown in Figure 19.

**Figure 19:**
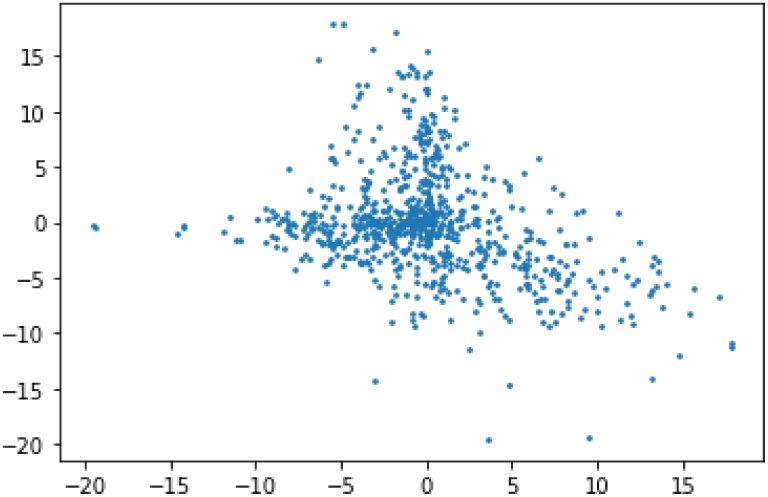
Phase space portrait of the residuals’ component.

**Figure 20:**
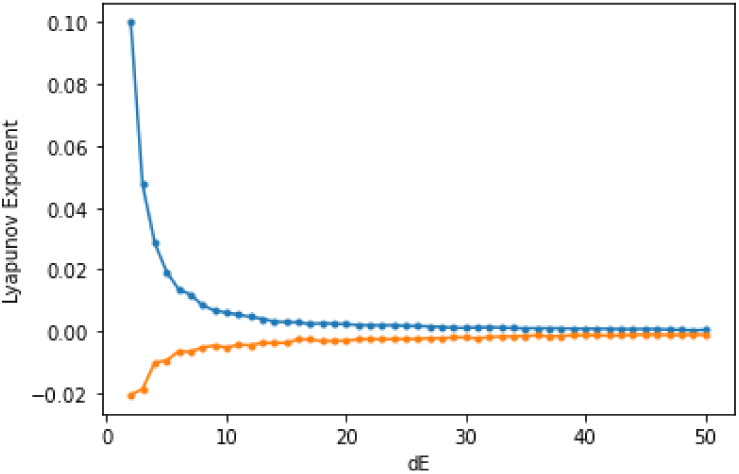
Lyapunov spectrum estimated for the residuals’ phase space dynamics.

Calculating the Lyapunov spectrum, we also find a convergence of the spectrum with a positive exponent given by 4.7035E-04 and a negative by -1.1240E-03, leading to a Kaplan-Yorke dimension of 1.4185.

Therefore, the evidence is also favorable to a weakly chaotic dynamics associated with the residuals. The issue is whether this dynamics is a second chaotic attractor or whether we are dealing with a stochastic process that is driven by the main chaotic attractor captured by the adaptive topological agent. It is this last hypothesis that we need to evaluate.

In this case we found that, upon testing multiple models, in order to remove all of the deterministic components, the actual dynamics for the residuals is comprised of a linear dependence upon the adaptive topological agent’s predictions, accounting for the chaotic signatures found above, a two-lag autoregressive dependence, which accounts for short-term recurrences found in the previous topological analysis and a GARCH component accounting for the nonstationary variance and turbulence markers.

Regarding the GARCH model type we found that there is a statistically significant mean dependence upon the dynamical variance, which leads to a GARCH in mean model. Formally, expanding from equation (10), the final model estimated for the residuals is given by the following equations:

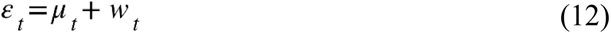

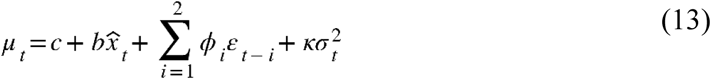

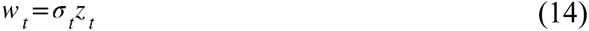

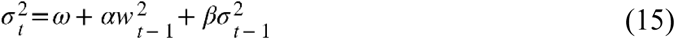

The above equations decompose the residuals’ series into a time-varying mean and a stochastic term *w* with zero mean and time-varying variance.

The time-varying variance is captured through the multiplication of a volatility parameter by an IID noise term *z* with zero mean and standard deviation of 1, as per equation (14), with the square of the volatility parameter, which is equal to the variance of the stochastic component *w*, having a dependence upon the square value of *w* and upon its previous value, which introduces an autoregressive dependence of the variance, this is the basic GARCH component, as per equation (15).

As per equation (13), the time-varying mean is given by a constant, a linear term that depends upon the adaptive topological agent’s predictions for that week, a two-lag autoregressive component and a term that depends upon the time-varying variance, which corresponds to the GARCH in mean model, which was first introduced in financial volatility modeling [27].

We also found that the distribution for *z* deviated from the normal distribution following a standardized Student’s *t* distribution, therefore, we used this last distribution for the final log-likelihood estimation.

As results from table 3, the coefficient directly associated with the adaptive topological agent’s term is statistically significant as are the coefficients associated with the residuals’ linear autoregressive component and the coefficient associated with the dynamical squared volatility, this means that the mean dependence upon the chaotic signal holds confirming the hypothesis that the signatures of chaos in the residuals’ dynamics are actually linked to the main series’ chaotic dynamics, captured by the adaptive topological agent.

**Table 3:** Estimation of the residuals’ model with dependence upon the adaptive topological agent’s predictions.

|  | Coef | std. err. | t | P>\|t\| | 95.0% Conf. Int. |
| --- | --- | --- | --- | --- | --- |
| *c* | -0.0093 | 0.0109 | -0.8550 | 0.3920 | [-3.054e-02,1.198e-02] |
| $\phi_1$ | 1.2737 | 0.0480 | 26.5180 | 0.0 | [ 1.180, 1.368] |
| $\phi_2$ | -0.4204 | 0.0401 | -10.4930 | 0.0 | [ -0.499, -0.342] |
| *b* | -0.0515 | 0.0067 | -7.7260 | 0.0 | [-6.460e-02,-3.846e-02] |
| $\kappa$ | 0.3831 | 0.0595 | 6.4400 | 0.0 | [ 0.266, 0.500] |
| $\omega$ | 0.0023 | 0.0017 | 1.3420 | 0.1800 | [-1.038e-03,5.546e-03] |
| $\alpha$ | 0.3624 | 0.0322 | 11.2410 | 0.0 | [0.299,0.426] |
| $\beta$ | 0.6376 | 0.0364 | 17.5120 | 0.0 | [0.566,0.709] |
| $\nu$ | 7.2354 | 0.9720 | 7.4440 | 0.0 | [5.331,9.140] |
| Metrics | Log Likelihood | R<sup>2</sup> | Adj. R<sup>2</sup> | AIC | BIC |
| Values | -1131.59 | 0.869 | 0.868 | 2281.18 | 2323.59 |

The negative value of the coefficient associated with the chaotic signal indicates that the residuals’ dynamics has a dampening effect upon the chaotic dynamics while at the same time it introduces a linear dependence upon the previous week, adding a weekly recurrence.

On the other hand, this positive dependence upon the previous week is reduced by a second lag weekly dependence. The two-lag structure explain the short range recurrences added by the residuals’ to the final attractor’s topological dynamics.

Finally, there is a mean positive dependence upon the nonstationary variance which confirms the GARCH in mean effect. The dominant component in the GARCH dynamics is the autoregressive dependence of the variance which leads to volatility buildups with impact on the mean of the process, through the GARCH in mean dependence, and adding further to the recurrence structure.

The coefficient of determination of the model is 86.90% and the adjusted coefficient of determination is 86.80%, which indicates that the residuals’ dynamics is captured well by the model.

As shown in Table 4, for the standardized noise term *z*, the null hypothesis of Engle’s LM and F tests is not rejected indicating that all of the ARCH effects are captured by the model.

**Table 4:** Engle’s tests and residuals’ distribution tests for the standardized noise term *z*.

|  | Statistic | p-value |
| --- | --- | --- |
| Engle's Test (LM) | 30.44973 | 0.2079 |
| Engle's Test (F) | 1.22505 | 0.2069 |
| Jarque-Bera Test | 49.67799 | 0.0 |
| Kolmogorov-Smirnov Test Normal | 0.045062 | 0.0686 |
| Kolmogorov-Smirnov Test Student's $t$ | 0.031617 | 0.3753 |

The Jarque-Bera’s test of normality leads to the rejection of the normality of *z* at a 1% level and the Kolmogorov-Smirnov test leads to the rejection of the normality at a 10% level, however, not for the Student’s *t* distribution, which shows that the Student’s *t* distribution is more appropriate for modeling the probabilistic dynamics of the noise component *z*, which was why we used it for the log-likelihood estimation, after a first attempt at using the Gaussian distribution which also led to the rejection of normality.

While Table 4’s results show, from Engle’s tests, that the nonlinearity associated with the GARCH component is captured by the model, the IID noise hypothesis still needs to be tested. From table 5, which presents the BDS test for dimensions from 1 to 10, we find that we cannot reject the BDS test’s null hypothesis of IID for the standardized noise term *z*.

**Table 5:** BDS test for the standardized noise term *z*.

| Embedding Dimension | Statistic | p-value |
| --- | --- | --- |
| 2 | 0.268687 | 0.788171 |
| 3 | -0.10795 | 0.914037 |
| 4 | -0.563 | 0.573437 |
| 5 | -0.58235 | 0.560329 |
| 6 | -0.49875 | 0.617958 |
| 7 | -0.76561 | 0.443909 |
| 8 | -0.64508 | 0.518874 |
| 9 | -0.42604 | 0.67008 |
| 10 | -0.41863 | 0.675485 |

In this way, we captured all of the deterministic pattern in the data, down to IID noise in the standardized noise process *z*. Confirming, also that we are dealing with stochastic chaos.

Using equation (10) and equations (12) to (15) along with table 3’s results, the following two equations hold for the main series’ stochastic process:

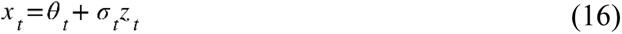

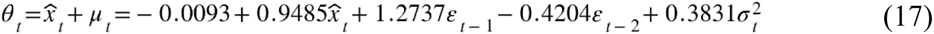

Equation (17) provides for the expected value of the final chaotically-driven stochastic process. This expected value is given by a constant, a term that depends upon the adaptive topological agent’s predictions that captured the chaotic signal, two linear autoregressive components and the GARCH in mean component.

The expected value calculated in equation (17) has an *R*^2^ of 97.14% in regards to the adjustment to the weekly share of positive tests, that is, it accounts for 97.14% of the variability of the epidemiological series, it also provides for an explained variance of 97.49%, and has a positive and strong correlation of 0.9875 with the main target series, an RMSE of 1.7104 which corresponds to 4.06% of the data amplitude, as per table 6.

**Table 6:** Main metrics for equation (17) expected value in relation to the weekly share of positive tests.

|  |  |
| --- | --- |
| R2 Score | 97.14% |
| Explained Variance | 97.49% |
| Correlation | 0.9875 |
| RMSE | 1.7103 |
| RMSE/Amp. | 4.06% |

In Figure 21, we show the weekly share of positive tests in blue and equation (17)’s expected value component dynamics in orange, as can be seen the main dynamics is closely matched by the time-varying expected value.

**Figure 21:**
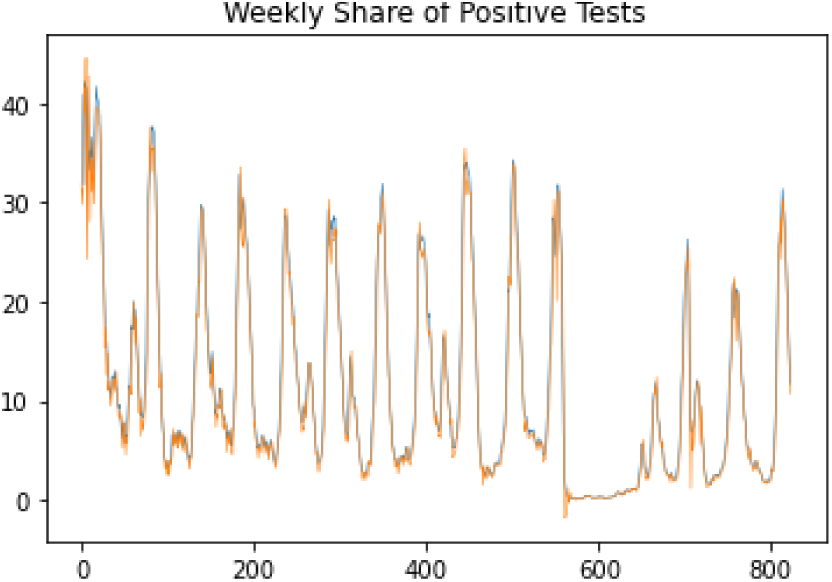
Dynamical expected value from equation (17) that includes the adaptive topological agent’s predictions plus the autoregressive dependence and GARCH in mean effect (orange) and observed values (blue) for the target epidemiological series.

Using the above results, we can also estimate a time series for the probability of the rate of positive tests to influenza surpassing a certain threshold, in this way, we can use this measure as an outbreak risk metric.

Indeed, using equations (12) to (17) and the standardized Student’s *t* distribution, we can calculate, for each time step, the probability of the level of positive tests for influenza to surpass a threshold *l* as follows:

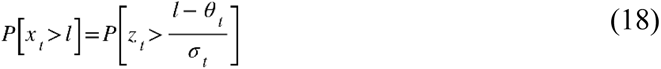

It is important to stress that, since the target epidemiological series is the rate of positive tests; the probability that is being calculated is the probability of that rate surpassing a specific threshold of positive tests, therefore, we are dealing with an influenza detection probability.

Estimating the above probability for different values of the threshold *l*, we find that the probability values change with time from low to high values, leading to clustering of the probability dynamics between periods of low probability and high probability, as can be seen in Figure 22, which shows the time series for the probability values and the corresponding recurrence plots, for threshold levels 5, 10, 15 and 20, respectively.

**Figure 22:**
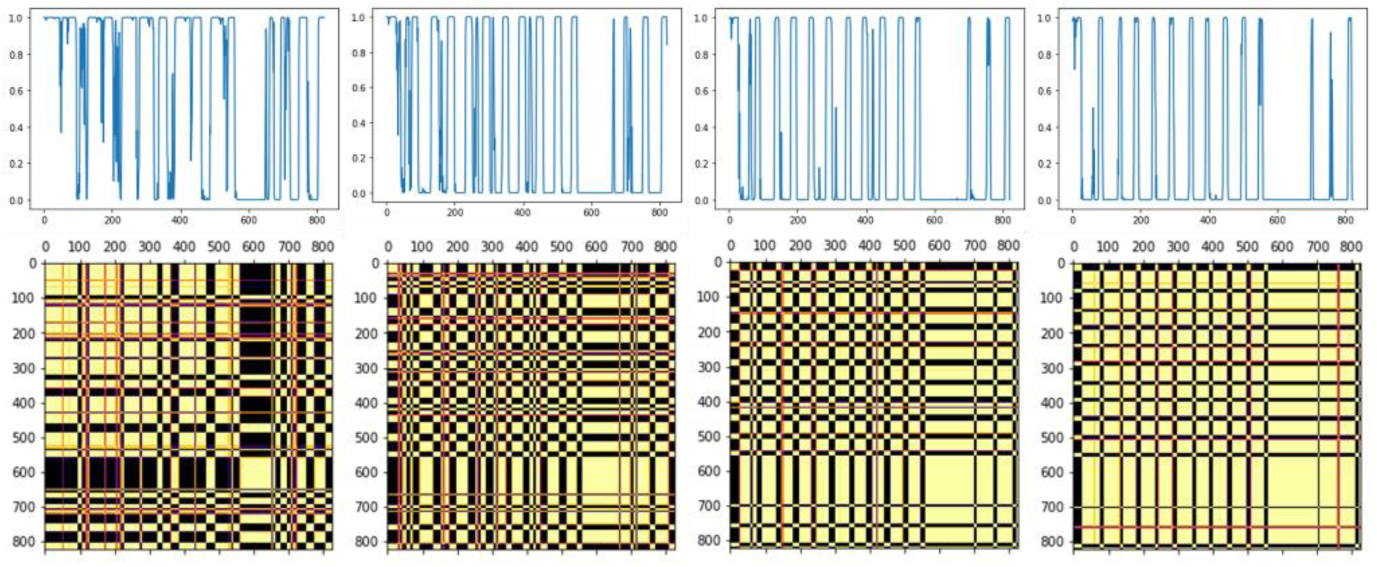
Probability dynamics (top) and corresponding colored recurrence plots (bottom) for *l* = 5, 10, 15, 20, respectively, Python’s Matplotlib’s reverse inferno colormap (“inferno_r”) was used.

The probability dynamics is not periodic but exhibits strong recurrences and intermittency. Indeed, as shown in Table 7, for a radius of 0.1, the recurrence probability is high for all of the analyzed thresholds, and the average recurrence strength is higher for the higher thresholds, which is linked to the intermittent dynamics associated with the outbreaks leading to longer periods of low detection probability interrupted by sudden rises in detection probability, which is linked to outbreaks.

**Table 7:** Recurrence metrics for equation (15)’s dynamical probability for the thresholds 5, 10, 15 and 20, respectively, using a radius of 0.1.

| | $l=5$ | $l=10$ | $l=15$ | $l=20$ |
| --- | --- | --- | --- | --- |
| % Recurrence | 100.00% | 99.88% | 99.76% | 99.51% |
| Average recurrence strength | 0.4143 | 0.4097 | 0.5187 | 0.5918 |
| P[100% recurrence\|recurrence] | 0.0219 | 0 | 0 | 0 |

By contrast, the conditional 100% recurrence probability drops with the rise of the threshold. This is also due to the outbreak dynamics, which leads to spikes in higher contagion events with sudden rises in probability interrupting the periods of lower contagion and breaking the 100% recurrence lines.

We find, in the recurrence plot, the impact of the COVID-19 event more clearly for the larger than for the smaller thresholds. This is due to the fact that a large influenza outbreak detection became less probable during the COVID-19 period, which was strongly due to the lockdowns and to the epidemiological prevalence of SARS-CoV-2 over influenza viruses during that period. This event had an impact on both the varying mean and the nonstationary variance dynamics.

Indeed, we find during the COVID-19 period a contraction of dispersion with a reduction of the dynamical volatility parameter, which has a direct impact both in the mean, through the GARCH in mean dependence, and the dispersion during this period.

In this way, while the COVID-19 period was consistent with the intermittent dynamics and did not lead to a deviation from the overall chaotic attractor dynamics, as captured by the adaptive topological agent, it effectively led to a volatility squeezing effect at the stochastic process level with impact in the residuals’ dynamics.

The process is captured well by the expected value as shown in Figure 21, which shows the low values of the rate of positive influenza tests during the COVID-19 period. We can also see these dynamics in the GARCH stochastic component *w* (equation 14) and the volatility dynamics (equation 15). Indeed, while in the standardized stochastic component *z* we do not find that squeezing effect, we do find it in the GARCH stochastic component *w* through the dynamical standard deviation, as can be seen in Figure 23.

**Figure 23.**
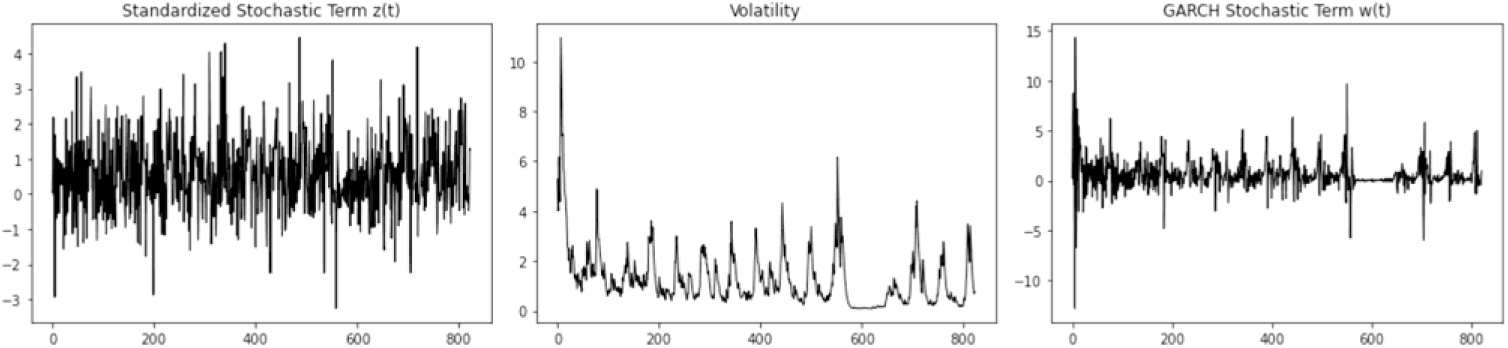
Standardized stochastic term (left), dynamical volatility parameter measured by the square root of the dynamical variance (middle) and GARCH stochastic component *w* (right).

Thus, the full joint chaotic and stochastic dynamics for the epidemiological series reflect the overall dynamics including during the COVID-19 period with high explanatory power.

## 3. Conclusion

Stochastic chaos is an open system’s dynamics that involves deterministic chaotic and stochastic components. Combining machine learning, chaos theory, and topological data analysis allows the development of topological robust methods for finding phase space embedding parameters for which the reconstructed attractors affected by dynamical noise have the highest exploitable topological markers for predicting a target series by an adaptive agent using a sliding window for relearning and a *k*-nearest neighbors’ learning unit.

By employing such an adaptive topological agent and testing the prediction performance on different embedding dimensions and delays, one is capable of finding embeddings of a time series where there is a stronger topological structure for the noisy reconstructed attractor linking that attractor to the target series, for which one can research the main topological properties for the reconstructed attractor.

Calculating the residuals from the adaptive topological agent’s predictions, one can further study the stochastic process down to the IID noise component, fully accounting for the deterministic and stochastic dynamics.

The application of this methodology to the weekly share of positive tests to influenza in the Northern Hemisphere allowed us to uncover a form of stochastic chaos down to the IID noise component, with a few relevant epidemiological findings.

First, there is evidence of stochastic chaotic dynamics with a two-dimensional chaotic attractor affected by a nonlinear stochastic process with nonstationary volatility captured by a GARCH in mean component. While the phase space is two-dimensional the attractor has a fractal structure that is between 1 and 2 dimensions but closer to 2, values that are obtained both from the Kaplan-Yorke and the box-counting dimension estimations.

The chaotic dynamics is linked to the formation of intermittent epidemiological contagion waves and is characterized by a power law scaling with a black noise spectrum, therefore to a strongly persistent dynamics that effectively compensates for the exponential amplification of small fluctuations linked to chaos, while at the same time it leads to long range impact of these fluctuations through a fractal memory. The testing process itself along with the underlying epidemiological outbreak dynamics, may be reinforcing the persistence of this process by leading to a possible undercounting of the actual number of positive cases, producing longer periods of low detection.

The result is that the attractor, while chaotic, is very close to the onset of chaos, with the low contagion periods leading to fixed-point-like signatures, which explains why the Kaplan-Yorke dimension is close to 2. The attractor profile further situates the chaotic dynamics into a form of weakly dissipative system, which implies an epidemiological model based on Hamiltonian chaos affected by a weakly dissipative component and by dynamical noise. This finding reinforces research directions studying Hamiltonian models with dissipative and stochastic components in epidemiology [28–30].

By analyzing the residuals from the adaptive topological agent’s predictions, which captured the chaotic signal, we were able to extract the full nonlinear stochastic chaotic process for the weekly rates of positive tests for influenza in the Northern Hemisphere as being comprised of a Student’s *t* distribution with a time-varying mean and standard deviation.

The standard deviation is described by a GARCH model, the time-varying mean is comprised of a constant plus a linear dependence on the chaotic signal captured by the adaptive topological agent’s predictions, plus two linear autoregressive dependence terms upon the series itself, the dominant term which expresses a dependence upon the previous week is positive but there is a second term that shows a linear dependence on a two week lag is negative. Finally, there is a dependence upon the variance of the process itself, which has a GARCH structure.

This leads to turbulence and volatility clustering associated with the epidemiological waves, with volatility waves associated with the outbreaks themselves. The epidemiological waves have an expression in both the chaotic dynamics, which is captured in the mean of the stochastic process, and the volatility process itself, which also impacts the mean through the dependence upon the dynamical variance.

Epidemiological events leave a direct mark in both the mean and the changing volatility measured by the standard deviation, we see this in particular in the COVID-19 period.

By modeling the stochastic chaotic dynamics down to IID noise we were also able to calculate outbreak probabilities and apply topological analysis methods to their dynamics.

The phase space decomposition showed that the chaotic attractor is structurally resilient to the stochastic component, with the main topological markers obtained for the embedding of the original series being close to those of the component captured by the adaptive topological agent.

Further studies are needed to study stochastic chaotic dynamics in influenza and other epidemiological series; namely, while Northern or Southern Hemisphere analysis may provide for aggregate macro-level results, the division into influenza transmission zones may provide for further insights into the underlying epidemiological dynamics.

## Data Availability

All data produced are available online at: https://ourworldindata.org/influenza

https://ourworldindata.org/explorers/influenza?tab=chart&country=~Northern+Hemisphere&Confirmed+cases+or+Symptoms=Confirmed+cases&Metric=Share+of+positive+tests+%28%25%29&Interval=Monthly&Surveillance+type=All+types

https://ourworldindata.org/influenza

## References

1. Oluwole O. Deterministic Chaos, El Niño Southern Oscillation, and Seasonal Influenza Epidemics. Front Environ Sci. 2017;5:8. doi:10.3389/fenvs.2017.00008.

2. Mangiarotti S, Peyre M, Zhang Y, et al.. Chaos theory applied to the outbreak of COVID-19: an ancillary approach to decision making in pandemic context. Epidemiology and Infection 2020;148, e95:1–9. doi:10.1017/S0950268820000990.

3. Postavaru O, Anton SR and Toma A. COVID-19 pandemic and chaos theory. Math. and Comp. in Sim. 2021;181:138–149. doi:10.1016/j.matcom.2020.09.029

4. Huang, YJ, Huang HT, Juang J., et al. Multistability of a Two-Dimensional Map Arising in an Influenza Model. J Nonlinear Sci 2022;32,15. doi:10.1007/s00332-021-09776-4.

5. Gonçalves CP. Low Dimensional Chaotic Attractors in SARSCoV-2’s Regional Epidemiological Data. Int J Swarm Evol Comput. 2022;11(9):1000271. doi: 10.35248/2090-4908.22.11.271.

6. Gonçalves CP. Low Dimensional Chaotic Attractors in Daily Hospital Occupancy from COVID-19 in the USA and Canada. Int J Swarm Evol Comput. 2023;11:291. doi: 10.35248/2090-4908.22.11.291.

7. Yılmaz E, Aydıner E. Chaotic and quasi-periodic regimes in the COVID-19 mortality data. Chaos Theory and Appl. 2024;6(1):41–50. doi: 10.51537/chaos.1420724.

8. Gonçalves CP. Epidemiological Rogue Waves and Chaos-Induced Multifractal Self-Organized Criticality in COVID-19. Int J Swarm Evol Comput. 2024;13:367. doi:10.35248/2090-4908.24.13.367.

9. Calistri A, Roggero PF, Palù G. Chaos theory in the understanding of COVID-19 pandemic dynamics. Gene. 2024;912:148334. doi:10.1016/j.gene.2024.148334.

10. Wagner J, Bauer S, Contreras S, et al.. Societal self-regulation induces complex infection dynamics and chaos. Phys Rev Res. 2025;7:013308. doi:10.1103/PhysRevResearch.7.013308.

11. Kaplan D, Glass L. Understanding Nonlinear Dynamics. Springer-Verlag, 1995. doi:10.1007/978-1-4612-0823-5.

12. Chen P. A Random Walk or Color Chaos on the Stock Market? Time-Frequency Analysis of S&P Indexes. Stud. Nonlinear Dyn. Econom. 1996;1(2):87–103. doi: 10.2202/1558-3708.1014.

13. Gonçalves CP. Topological Machine Learning and Chaotic Attractors Decomposition–An Application to Sunspot Chaos. Int J Swarm Evol Comput. 2024;13:387. doi:10.35248/2090-4908.24.13.387.

14. Gonçalves CP. Stochastic Chaotic Network Vector Fields. Int J Swarm Evol Comput. 2025;14:393. doi:10.35248/2090-4908.25.14.393.

15. Frey M, Simiu E. Noise-induced chaos and phase space flux. Phys D 1993;63:321–340. doi:10.1016/0167-2789(93)90114-G.

16. Kaneko K, Tsuda I. Complex Systems: Chaos and Beyond: Chaos and Beyond: A Constructive Approach With Applications in Life Sciences. Springer Science & Business Media. 2001. doi:10.1007/978-3-642-56861-9.

17. Cvitanović P, Artuso R, Mainieri R, et al. Chaos: Classical and quantum [Internet]. Copenhagen: Niels Bohr Institute; 2023 [cited 2025 May 1]. Available from: https://chaosbook.org/.

18. Eckmann JP, Kamphorst SO, Ruelle D. Recurrence plots of dynamical systems. Europhys Lett. 1987;4:973–977. doi:10.1209/0295-5075/4/9/004.

19. Takens F. Detecting strange attractors in turbulence. In: Rand DA, Young LS, editors. Dynamical systems and turbulence. Lecture Notes in Mathematics. Vol. 898. Berlin: Springer-Verlag; 1981. p. 366–81. doi:10.1007/BFb0091924.

20. Eckmann JP, Kamphorst SO, Ruelle D, Ciliberto S. Liapunov exponents from time series. Phys Rev A. 1986 Dec;34(6):4971–4979. doi: 10.1103/PhysRevA.34.4971.

21. Kaplan JL, Yorke JA. Chaotic behavior of multidimensional difference equations. In: Peitgen HO, Walther HO, editors. Functional Differential Equations and Approximation of Fixed Points. Lecture Notes in Mathematics. Vol. 730. Berlin: Springer; 1979. p. 204–227. doi:10.1007/BFb0064319.

22. Peitgen HO, Jürgens H, Saupe D. Chaos and Fractals: New Frontiers of Science. 2nd ed. Springer; 2004. doi: 10.1007/b97624.

23. Haken H. Synergetics: Introduction and Advanced Topics. Berlin: Springer-Verlag; 2004. doi: 10.1007/978-3-662-10184-1.

24. Brock WA, Dechert WD, Scheinkman JA. A test for independence based on the correlation dimension. Department of Economics, University of Wisconsin, Madison; 1987. SSRI Working Paper No. 8702.

25. Engle RF. Autoregressive conditional heteroskedasticity with estimates of the variance of United Kingdom inflation. Econometrica. 1982;50(4):987–1007. doi:10.2307/1912773

26. Schroeder MR. Fractals, chaos, power laws: minutes from an infinite paradise. New York: W.H. Freeman; 1991.

27. Engle RF, Bollerslev T. GARCH in mean. Econometrica. 1987;55(4):775–790. doi: 10.2307/1911031.

28. Kamenev A, Meerson B. Extinction of an infectious disease: a large fluctuation in a non-equilibrium system. Phys Rev E. 2008;77(6):061107. doi: 10.1103/PhysRevE.77.061107.

29. Nakamura GM, Martinez AS. Hamiltonian dynamics of the SIS epidemic model with stochastic fluctuations. Sci Rep. 2019;9:15841. doi: 10.1038/s41598-019-52351-x.

30. Blázquez-Sanz D, Rodríguez M, Sardón C. Hamiltonian structure of compartmental epidemiological models. Physica D. 2020;413:132674. doi: 10.1016/j.physd.2020.132674.

